# Ruling In and Ruling Out Sepsis Using Likelihood Ratios of a Host Response Assay

**DOI:** 10.64898/2026.05.29.26354374

**Authors:** Krupa A. Navalkar, Prashant Wani, Roy F. Davis, Silvia Cermelli, Maximilian Dietrich, Maik von der Forst, Sören L. Becker, Sophia Benthien, Elisa Baumann, Carsten Zeiner, Philipp M. Lepper, José Garnacho-Montero, María Luisa Cantón-Bulnes, Adela Fernández-Galilea, Jose Luis García-Garmendia, Ángel Estella, Russell R. Miller, Marcus J. Schultz, Richard Rothman, John Burke, Gourang Patel, Jorge Parada, Thomas D. Yager, Richard B. Brandon

**Affiliations:** Immunexpress Inc., Seattle, WA 98109, USA; Heidelberg University, Medical Faculty Heidelberg, Department of Anesthesiology, Im Neuenheimer Feld 420, 69120 Heidelberg, Germany; Institute of Medical Microbiology and Hygiene, Saarland University, Homburg, Germany; Department of Internal Medicine V, Pneumology and Intensive Care Medicine, Saarland University, Homburg, Germany; Department of Internal Medicine, Pneumology and Intensive Care Medicine, University Hospital OWL Campus Bethel and University of Bielefeld, Bielefeld, Germany; Intensive Care Unit, Hospital Universitario Virgen del Rocío, Sevilla, Spain; Intensive Care Unit, Hospital Universitario Virgen Macarena, Sevilla, Spain; Intensive Care Unit, Hospital San Juan de Dios del Aljarafe, Sevilla, Spain; Intensive Care Unit, Hospital Universitario de Jerez de la Frontera, Cádiz, Spain; Instituto de Investigación e Innovación Biomédica de Cádiz (INiBICA), Department of Medicine and Surgery, University of Cádiz, Cádiz, Spain; FirstHealth of the Carolinas, Pinehurst, NC 28374, USA; Department of Anaesthesia, General Intensive Care and Pain Management, Division of Cardiothoracic and Vascular Anaesthesia & Critical Care Medicine, Medical University Wien, Vienna, Austria; Nuffield Department of Medicine, University of Oxford, Oxford, UK; Mahidol–Oxford Tropical Medicine Research Unit (MORU), Mahidol University, Bangkok, Thailand; School of Medicine, Johns Hopkins University, Baltimore, MD 21205, USA; Intermountain Medical Center, Murray, UT 84107, USA; School of Medicine, University of Utah, Salt Lake City, UT 84132, USA; University of Chicago Medicine, Chicago, IL 60637, USA; Loyola University Medical Center, Maywood, IL 60153, USA

**Keywords:** sepsis, SeptiCyte RAPID, likelihood ratio, host-response biomarker, systemic inflammatory response syndrome, SeptiScore, diagnostic accuracy, blood culture, intensive care unit, gene expression

## Abstract

**Overview:** SeptiCyte RAPID is an FDA-cleared gene expression test that quantifies host immune response to aid in the diagnosis of sepsis. The test yields a score (the SeptiScore) ranging from 0-15, distributed across four bands (1-4) based on increased likelihood of sepsis. Each band can be characterized by average positive and negative likelihood ratios (LR+, LR-respectively) for the discrimination of sepsis versus the non-infectious systemic inflammatory response syndrome (SIRS).

**Methods:** A retrospective analysis of prospectively collected data from a combined cohort of critically ill patients suspected of sepsis (N=889), recruited across 19 hospitals in the USA and Europe. The analysis quantified the LR+ and LR-parameters as a function of SeptiScore, for discrimination of sepsis vs. SIRS in patients admitted to ICU.

**Hypotheses:** (1) The likelihood ratio (LR) framework provides a clinically useful interpretive approach that complements the previously used SeptiScore banding scheme; (2) Low Band 1 SeptiScores are associated with sufficiently small LR-to support the use of SeptiCyte RAPID as a rule-out test for sepsis; (3) High Band 4 SeptiScores are associated with sufficiently large LR+ to support the use of SeptiCyte RAPID as a rule-in test for sepsis; and (4) SeptiScore-derived LR+ and LR-values can be combined with estimates of pre-test probability (derived from patient characteristics and/or other diagnostic tests) to generate individualized, patient-specific post-test probabilities of sepsis.

**Results:** The SeptiCyte RAPID test demonstrates strong diagnostic performance in distinguishing sepsis from SIRS. The likelihood ratios across different score bands provide clear clinical utility: the median LR+ was 3.26 (range 2.57-4.24) for Band 3, and 6.97 (range 4.35-15.57) for Band 4 providing evidence toward ruling in sepsis at high SeptiScores. Conversely, the median LR-was 0.16 (range 0.14-0.20) for Band 2 and 0.085 (range 0.014-0.16) for Band 1, providing evidence toward ruling out sepsis at low SeptiScores. A higher-resolution analysis of SeptiCyte RAPID performance confirmed these trends by evaluating LR+ and LR-at specific values within each band. The sepsis group was further stratified according to whether patients were classified as blood-culture positive (BC+) or blood culture negative (BC-), and the detailed LR+ and LR-analyses were repeated. A monotonic increase in likelihood ratio with increasing SeptiScore was consistently observed, independent of whether sepsis patients were culture-positive, culture-negative, or unstratified with respect to blood culture status.

**Conclusion:** High SeptiScores have correspondingly high LR+ values, and low SeptiScores have correspondingly low LR-values, both of which may have clinical utility. High likelihood ratios for band 4 SeptiScores, which precede traditional microbiology results, may provide clinicians with early confidence of a sepsis diagnosis and microbiology diagnostic stewardship. Low likelihood ratios for band 1 SeptiScores may prompt clinicians to consider an alternate diagnosis to sepsis. Such results, obtained early in the diagnostic workup process, may lead to fewer missed diagnoses and more efficient use of hospital resources.

## 1. Introduction

SeptiCyte RAPID is an FDA-cleared real-time quantitative RT-PCR test that measures the relative expression levels of two host immune response genes, PLA2G7 and PLAC8, which are relevant to the diagnosis of sepsis [1]. The test yields a SeptiScore, equal to the difference in threshold cycle (Cq) values between the PLA2G7 and PLAC8 amplicons. The SeptiScore ranges from 0 – 15, with higher SeptiScores indicating an increased likelihood of sepsis. The SeptiScore range has been divided into four Bands (B1 SeptiScores 0-4.9; B2 SeptiScores 5.0-6.1; B3 SeptiScores 6.2-7.3; B4 SeptiScores 7.4-15, Supplement S1) [1]. Each SeptiScore Band can be characterized by performance measures that are significantly affected by sepsis prevalence such as negative and positive predictive values. SeptiScore Bands can also be characterized by prevalence-independent measures such as sensitivity, specificity, and band-specific likelihood ratios (LR+ and LR-).

The likelihood ratio (LR) concept provides a context-dependent framework for evaluating SeptiCyte performance by combining both sensitivity and specificity into a single, stable measure that quantifies how much a test result changes the likelihood of disease [2]. Unlike other performance measures which describe test accuracy in isolation, LRs usefully integrate pre-test and post-test probabilities through Bayes’ theorem, thereby offering a direct measure of how test results can modify a clinician’s *a priori* estimate of disease probability. The use of LRs thus may enhance clinical decision-making by translating statistical accuracy into meaningful disease probability estimates that are relevant to patient care [3].

This study describes a detailed LR analysis for SeptiCyte RAPID in a combined cohort of critically ill (ICU) patients suspected of sepsis (N=889) recruited in 19 hospitals across the USA and Europe. Besides estimating LR+ and LR-for each SeptiScore band, a higher-resolution analysis of SeptiCyte RAPID performance was also considered, in which the LR+ and LR-were evaluated across a continuous scale of SeptiScore values, rather than being restricted to the four pre-defined interpretation bands. A further stratification was also conducted, in which the sepsis patient group was stratified with respect to being blood culture positive (BC+) or blood culture negative (BC-), and the LR analysis repeated.

Four hypotheses are addressed in this study: (1) that the likelihood ratio (LR) framework provides a clinically useful interpretive approach that complements the previously used SeptiScore banding scheme; (2) that low Band 1 SeptiScores are associated with sufficiently small negative likelihood ratios (LR-) to support the use of SeptiCyte RAPID as a rule-out test for sepsis; (3) that high Band 4 SeptiScores are associated with sufficiently large positive likelihood ratios (LR+) to support the use of SeptiCyte RAPID as a rule-in test for sepsis; and (4) that SeptiScore-derived LR+ and LR-values can be combined with estimates of pre-test probability (derived from patient characteristics and/or other diagnostic tests) to generate individualized, patient-specific post-test probabilities of sepsis.

## 2. Materials and Methods

### 2.1. Study Cohorts

Figure 1. presents a consort diagram describing the origins of the patients in this study. The patients were recruited from ICUs across 19 sites in Europe and the United States. Details of the individual patient cohorts in this study, including descriptions of all patient exclusions are presented below.

**Figure 1.**
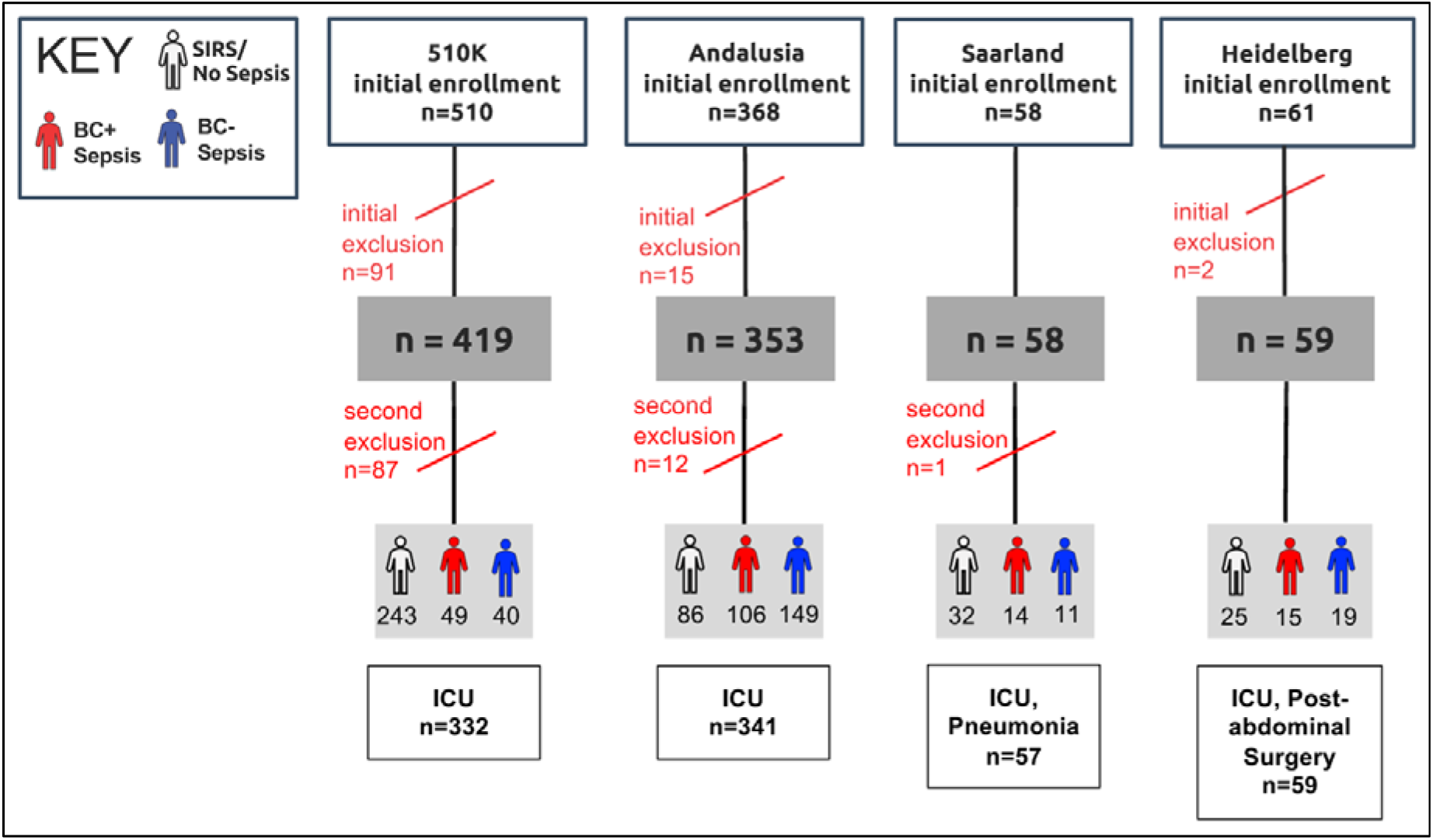
Consort diagram for sepsis vs SIRS discrimination. Four cohorts were recruited across 19 sites. Total all sepsis = 503. Total BC(+) sepsis = 184. Total BC(−) sepsis = 219. Total SIRS = 386. Total for sepsis/SIRS analysis without BC stratification = 889. Total for sepsis/SIRS analysis with BC(+/-) stratification = 789.

#### Immunexpress 510k cohort (final N = 419)

This cohort consisted of retrospective (N = 356) and prospective (N = 63) components. A full description of the study cohort and a flow diagram for the origin of all samples used in the study has previously been published [1]. The retrospective component of critically ill (ICU) patients was drawn from the observational MARS, VENUS, and VENUS Supplement trials (NCT01905033 and NCT02127502 on clinicaltrials.gov). The recruitment dates were January 2011–December 2013 (MARS), May 2014–April 2015 (VENUS) and March–August 2016 (VENUS Supplement). The prospective component consisted of 63 critically ill (ICU) adult subjects enrolled in an observational trial (NEPTUNE, NCT05469048; clinicaltrials.gov (accessed on 17 February 2024)) between the dates 26 May 2020 and 25 April 2021 at Emory University/Grady Memorial Hospital (Atlanta, GA, USA), Rush University Medical Center (Chicago, IL, USA) and University of Southern California (USC) Medical Center (with two separate sites, Keck Hospital of USC, and Los Angeles General Medical Center, in Los Angeles, CA, USA). The NEPTUNE inclusion/exclusion criteria matched the criteria for the earlier studies.

*initial enrolment:* 510 ICU patients

*first-stage exclusion:* 91 excluded because banked PAXgene blood sample no longer available

*second-stage exclusion:* 87 sepsis patients excluded because no BC (+/-) results documented

*N for unstratified analysis:* 419

*N for BC+/− analysis:* 332

*243 SIRS*

*49 BC(+) sepsis*

*40 BC(−) sepsis*

##### Comparator

Classification as sepsis or SIRS was achieved through a process of retrospective physician diagnosis (RPD) by a panel of three expert clinicians not involved with the care of the patients. The method has been thoroughly described in Supplement 3 of Miller et al. (2018) [4]. If at least two out of the three panelists agreed that the patient had sepsis or SIRS, then that adjudication was accepted as Consensus RPD, leaving 41 (9.8%) indeterminate cases. Patients with an “indeterminate” call underwent an additional independent blinded case review and forced into a binary classification of sepsis or SIRS. Under this secondary adjudication (“Forced RPD”) no indeterminate calls were allowed, lending access to the full cohort.

#### Heidelberg cohort (N=59)

The “SeptAsTERS” study was conducted at a single hospital in Germany. The final enrollment was of 59 post-surgical ICU patients with SIRS and showing signs of clinical deterioration. A total of 34 (57.6%) were retrospectively determined to have sepsis, and the remaining 25 (42.4%) were determined to have SIRS. Blood cultures were ordered on the basis of clinical judgement and accordingly were only taken if deemed necessary by the care team. Of the 34 septic patients, 15 (44.1%) were blood culture positive.

*Initial enrolment:* 61 ICU patients

*first-stage exclusion:* 2 patients excluded (1 readmission, and 1 palliative care) *second-stage exclusion:* none

*N for unstratified analysis:* 59 *N for BC+/− analysis:* 59

*25 SIRS*

*15 BC(+) sepsis*

*19 BC(−) sepsis*

##### Comparator

The comparator for classification as SIRS or sepsis in this study was a post-hoc assessment of all patients by three independent intensive care professionals, who were not involved in the study. The process followed a model similar to the one described in the online data supplement Part 3 by Miller et al. (2018) [4] based on FDA Guidance and on publications by Klein Klouwenberg et al (2013) [5]. Details of the post-hoc assessment process are provided in von der Forst et al. (2024) [6]. As organ dysfunction parameters were considered, the assessment can be viewed as having been performed under the Sepsis-3 conceptual framework [7].

#### Andalusia cohort (N=353)

(Cantón-Bulnes et al., 2026) [8]: An observational, prospective, and multicenter study was conducted between March 3, 2022 and December 20, 2022 in seven ICUs in Andalusia (Spain) and coordinated by the Virgen Macarena University Hospital in Seville. A total of 354 patients were enrolled, of whom 86 (24.3%) did not present sepsis at the researchers’ discretion. All patients aged 18 years or older, admitted to the ICU, with a diagnosis of sepsis, according to the Sepsis-3 definition were included. Subjects were excluded if they were pregnant, or the clinical picture suggestive of sepsis had started more than 48 hours previously.

*initial enrolment:* 368 ICU patients

*first-stage exclusion:* 15 patients excluded (8 consent not given; 2 satisfying exclusion criteria; 3 with analytical failures; 1 duplicate; 1 for non-consensual classification) (see Supplemental

Figure 2 in Cantón-Bulnes et al., 2026)

**Figure 2.**
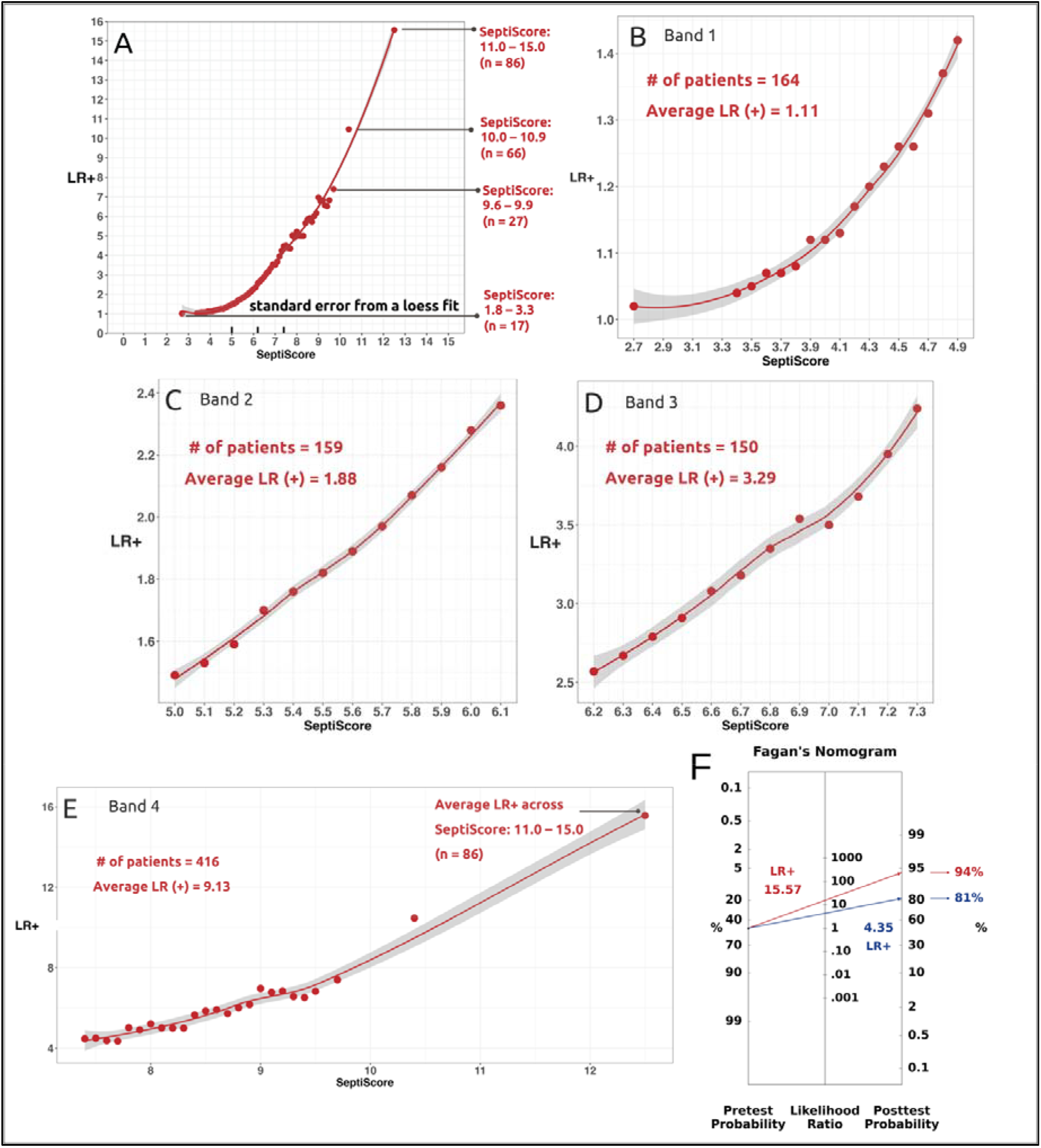
Positive likelihood ratio (LR+) analysis of complete dataset (N=889). No stratification by BC(+/-) was performed. **(A)** LR+ versus SeptiScore, over entire SeptiScore region. **(B)** Expanded view of Band 1 region. **(C)** Expanded view of Band 2 region. **(D)** Expanded view of Band 3 region. **(E)** Expanded view of Band 4 region. **(F)** Fagan nomogram for SeptiScore Band 4, assuming a pre-test sepsis prevalence of 50% and LR+ values of 4.35 and 15.57. (These LR+ values correspond to the Band 4 lower and upper bounds, respectively.)

*second-stage exclusion:* 12 sepsis patients excluded because no BC (+/-) results documented *N for unstratified analysis:* 353

*N for BC+/− analysis:* 341

*86 SIRS*

*106 BC(+) sepsis*

*149 BC(−) sepsis*

##### Comparator

In a retrospective analysis, patients were categorized as “sepsis” or “sterile inflammation” (i.e. SIRS) by two investigators at each participating site who were aware of the clinical, analytical, and microbiological data but blinded to the SeptiCyte results. A Steering Committee reviewed all cases and contacted the site investigators in case of doubts.

#### Saarland cohort (N=58) (Benthien et al. 2024) [9]

Patients aged ≥ 18 years at or submitted to the pneumological ICU with a suspicion of sepsis, constitute the eligible study patient population. Patients aged < 18 years, pregnant patients, as well as the absence of written informed consent constitute cases that are not be considered for study entry (exclusion criteria). In addition, patients who were on therapeutic antibiotic treatment for longer than 48 hrs prior to SeptiCyte RAPID sampling were also excluded.

Patients were recruited from the ICU at Pneumology and Intensive Care Medicine, Saarland University, Homburg, Germany. Between November 2022 and February 2024, a total of 39 symptomatic patients with a sepsis-like clinical pattern and 18 non-septic controls from the same ICU were recruited. Subjects were included if they had a change in the SOFA score within 24 hours of ICU admission of >=2 and antimicrobial therapy for <=48 hours. After having obtained informed consent, peripheral blood samples from the patients were collected using PAXgene RNA tubes and subjected to the SeptiCyte RAPID assay. Clinicians were blinded to the SeptiCyte results.

*initial enrolment:* 58 ICU patients

*first-stage exclusion*: none

*second-stage exclusion:* 1 sepsis patient excluded because no BC (+/−) results documented

*N for unstratified analysis:* 58

*N for BC+/− analysis:* 57

*32 “SIRS” (i*.*e. non-septic controls)*

*14 BC(+) sepsis*

*11 BC(−) sepsis*

##### Comparator

RPD was conducted with a panel of clinicians not involved in patient care. The RPD process led to a final clinical adjudication of 26 patients with infection-related sepsis, and 32 patients with non-sepsis status which are placed in the SIRS category.

### 2.2. Ethics Approval and Consent to Participate

All research reported in this study was conducted in accordance with the Declaration of Helsinki, and in accordance with relevant guidelines and regulations. Informed Consent was obtained from all subjects or their legal representatives.

#### Heidelberg

The SeptAsTERS component of this study was approved by the Ethics Committee of the Medical Faculty of Heidelberg University (S-118/2021) and was registered in the German Clinical Trials Register (DRKS00024891) prior to enrollment.

#### MARS

Ethics approval for the MARS trial was given by the Medical Ethics Committee of the Amsterdam Medical Canter (approval # 10-056C).

#### VENUS

Ethics approvals for the VENUS trial were given by the relevant Institutional Review Boards as follows: Intermountain Medical Center/Latter Day Saints Hospital (approval # 1024931); Johns Hopkins Hospital (approval # IRB00087839); Rush University Medical Center (approval # 15111104-IRB01); Loyola University Medical Center (approval # 208291); Northwell Healthcare (approval #16-02-42-03).

#### NEPTUNE

Ethics approvals for the NEPTUNE trial were given by the relevant Institutional Review Boards as follows: Emory University (approval # IRB00115400); Grady Memorial Hospital (approval # 00-115400); Rush University Medical Center (approval # 19101603-IRB01); University of Southern California Medical Center (approval # HS-19-0884-CR001).

#### Andalusia

The study was approved by the Research Ethics Committees of the Virgen Macarena-Virgen Rocio University Hospitals on 20 December 2021 (Internal Code, 2662-N-21). Since sepsis is a time-dependent process and the clinical outcome depends on how quickly it is recognized, this assay should be performed as soon as possible after clinical suspicion of sepsis. Based on this argument, the research ethics committees allowed blood sample extractions prior to obtaining written permission. Written consent from the patient or next of kin was obtained within 48 hours of ICU admission and the blood sample was discarded if written consent was not obtained. The study was approved by the Research Ethics Committees of the Virgen Macarena-Virgen Rocio University Hospitals (Certificate of Ethics Approval dated 23 Dec 2021. Trial name SEPT-ANC. Codigo Interno: 2662-N-21.)

#### Saarland

The study was approved by the Ethics Commission of the Saarland Medical Association for Saarland University (approval # 68/22, date 23 May 2022).

### 2.3. Statistical Methods

Calculations of the area under the receiver operating characteristic curve (AUC), sensitivity, and specificity were performed in R using the *pROC* package (version 1.19.0.1), specifically the roc function, as described by Robin et al. (2011) [10]. To estimate likelihood ratios (LRs) across the full range of SeptiScore values, two complementary approaches were used to account for differences in data density.

1. In the central region of the SeptiScore distribution, where observations were relatively abundant, LRs were derived from receiver operating characteristic (ROC) analysis using corresponding estimates of sensitivity and specificity at the relevant operating points:

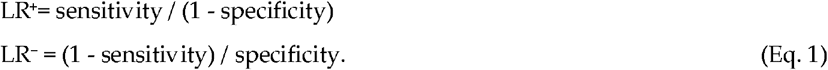
2. In the tails of the SeptiScore distribution where data were sparse, ROC-based estimates become unstable due to limited sample density. To improve statistical robustness in these tail regions, we binned SeptiScore values into intervals containing at least five patient observations per bin (≥5 samples per interval). Likelihood ratios (LR^+^ and LR^−^) were calculated empirically from the resulting 2×2 contingency tables constructed within each interval. The relationship between LR+, LR− and the contingency table values of true positives (TP), false positives (FP), true negatives (TN), and false negatives (FN) is given by the following derivation:

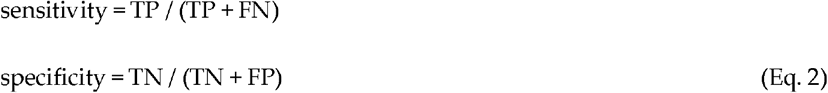

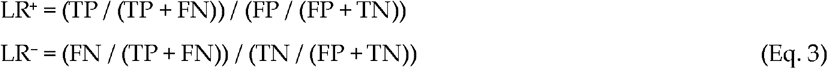

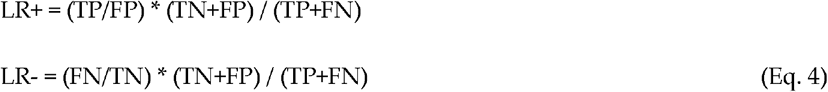

The positive and negative likelihood ratios (LR+ and LR-) were plotted as functions of SeptiScore. A smoothed best-fit curve was generated using local polynomial regression (LOESS) as implemented in the geom_smooth function of the R ggplot2 package (version 3.5.2). In figures below, the gray shaded band surrounding the LOESS curve represents the 95% confidence interval for the fitted relationship. This confidence interval is derived from the standard errors of the LOESS fit, assuming these errors follow an approximate Student’s t distribution.

As a visualization aid, we used a custom python program to construct Fagan nomograms in log-odds space to represent the relationship between pre-test probability, likelihood ratio (LR), and post-test probability. In this space, the post-test log-odds is the sum of the pre-test log-odds and the log of the LR. To ensure that lines connecting pre-test probabilities to post-test probabilities via the LR axis appear straight, the post-test axis was affine-transformed (both shifted and rescaled) relative to the pre-test axis. This adjustment preserves the additive property of log-odds and allows multiple LRs to be plotted simultaneously, providing an accurate visual representation of post-test probabilities [11].

## 3. Results

### 3.1. Patient Cohorts

We recruited a total of 889 intensive care patients comprising four cohorts from 19 hospital sites in Europe and USA. Data from 789 of the patients were available for the BC +/− analysis. In total, 184 BC(+) sepsis patients, 219 BC(−) sepsis patients, 100 sepsis patients without BC data, and 386 SIRS patients were included in the analyses. **Figure 1** in Materials and Methods presented a flow diagram showing patient numbers from each study that were included or excluded, and the number of patients retrospectively determined to be sepsis or SIRS, or BC+ / BC-sepsis. **Table 1** below summarizes some basic demographic data for these patient cohorts. Although the patients were all recruited from the ICU, considerable diversity is evident with respect to race/ethnicity, age and mortality rate.

**Table 1.**
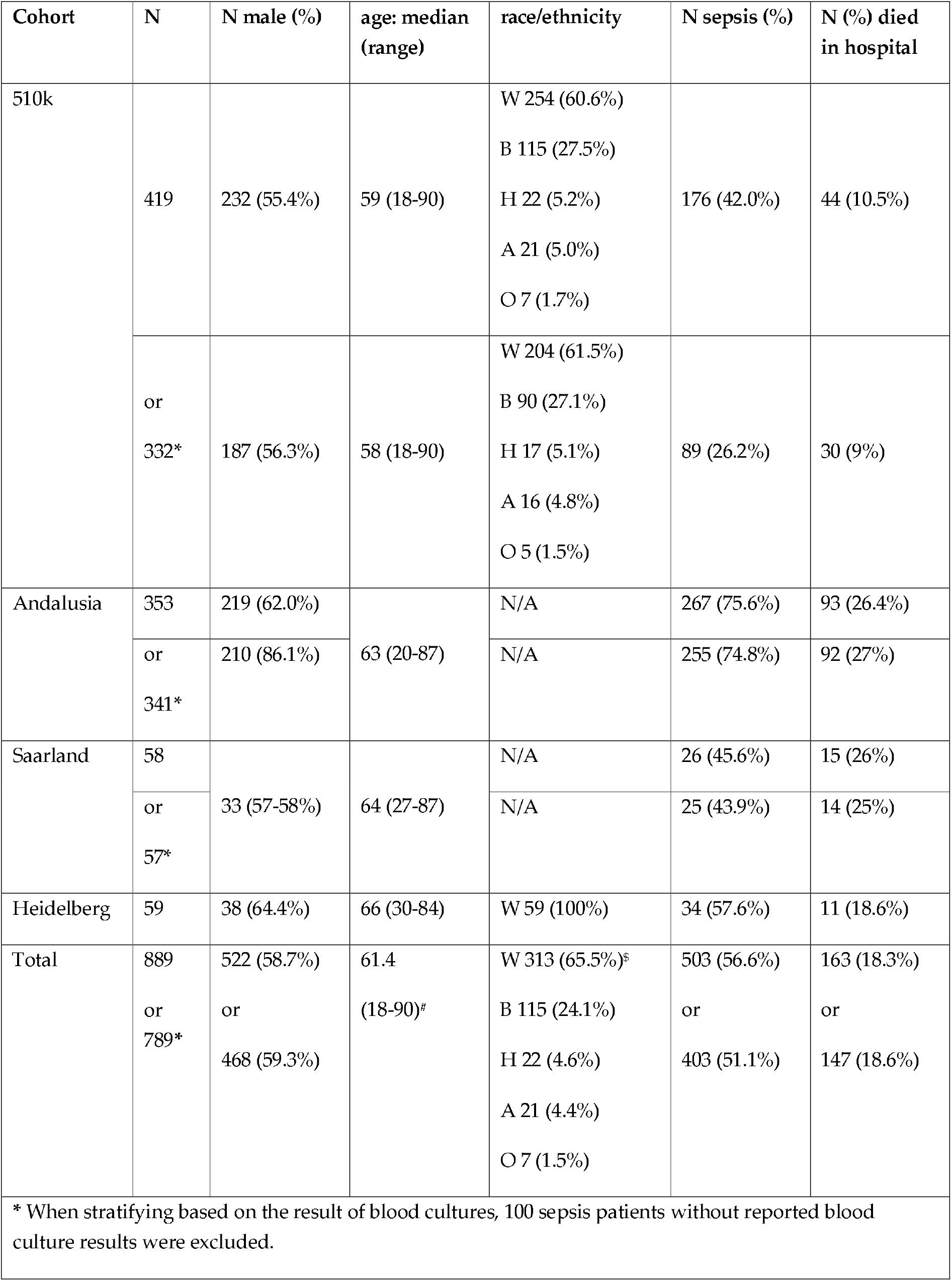
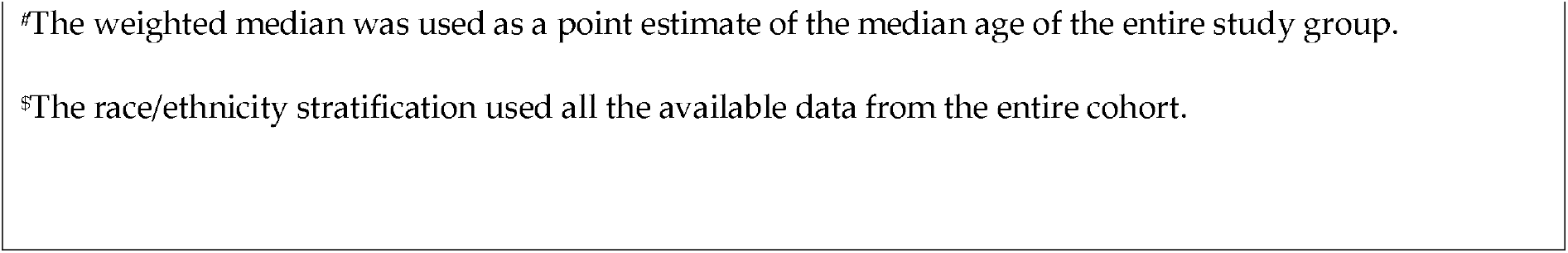
Demographics, including either all patients (N=889), or after excluding* sepsis patients for whom no BC results were available (N=789). Abbreviations: A (Asian), B (Black), H (Hispanic), W (White), O (other / unknown), N/A (data not available).

### 3.2. Positive Likelihood Ratios (LR+)

**Table 2** and **Figure 2** show, for the entire available dataset (4 cohorts, N=889) the relationship between the SeptiScore and the positive likelihood ratio (LR+). There is a clear monotonic increase in LR+ as a function of increasing SeptiScore.

**Table 2.** Positive Likelihood Ratios for sepsis/SIRS discrimination in the complete dataset (N=889). The minimum (min), maximum (max), median and mean LR+ values for each SeptiScore Band were estimated from the distributions provided in Supplement S2.

| SeptiScore Band | Range | LR+ min, or at lower boundary | median LR+ | mean LR+ | LR+ max, or at upper boundary | N sepsis | N SIRS | N total |
| --- | --- | --- | --- | --- | --- | --- | --- | --- |
| 1 | 0.0-4.9 | 1.00 | 1.07 | 1.11 | 1.42 | 25 | 139 | 164 |
| 2 | 5.0-6.1 | 1.49 | 1.86 | 1.88 | 2.36 | 42 | 117 | 159 |
| 3 | 6.2-7.3 | 2.57 | 3.26 | 3.29 | 4.24 | 81 | 69 | 150 |
| 4 | 7.4-15.0 | 4.35 | 6.97 | 9.13 | (average for SeptiScore 11-15: 15.57) | 355 | 61 | 416 |
| totals |  |  |  |  |  | 503 | 386 | 889 |

In Figure 2, **Panel A** shows a graph of LR+ versus SeptiScore over the entire SeptiScore range, for the complete dataset of 503 sepsis vs. 386 SIRS patients. **Panels B, C, D, E** show expanded views of the Band 1, 2, 3, 4 regions, respectively. In these panels the 95% uncertainty cloud around the LOESS best fit line is shown in grey. As an illustrative example, **Panel F** shows the Fagan nomogram corresponding to the low (4.35) and high (15.57) LR+ values for SeptiScore Band 4, assuming a pre-test sepsis probability of 50%. Note: in this and subsequent nomograms, the post-test probability axis has been affine transformed (shifted and stretched) relative to the pre-test probability and LR axes, so as to present the pre-test / post-test relationships as straight lines on this type of plot.

**Table 3** and **Figure 3** stratify the above analysis according to whether, in the comparison to SIRS, the septic patients could be classified as BC(+) or BC(−). (A total of 100 septic patients without reported BC data were excluded in this secondary analysis.)

**Table 3.** LR+ analysis per SeptiScore Band, stratified according to whether BC(+) sepsis or BC(−) sepsis is being compared to SIRS. For this secondary analysis, 789 patients were used. The minimum (min), maximum (max), median and mean LR+ values for each SeptiScore Band were estimated from the distributions provided in Supplements S3 and S4.

| SeptiScore Band | SeptiScore Range | median LR+ for BC+ sepsis vs SIRS | mean LR+ for BC+ sepsis vs SIRS | median LR+ for BC- sepsis vs SIRS | mean LR+ for BC- sepsis vs SIRS | No. of patients |
| --- | --- | --- | --- | --- | --- | --- |
| 1 | 0.0-4.9 | 1.07 | 1.13 | 1.07 | 1.11 | 153 |
| 2 | 5.0-6.1 | 1.97 | 1.99 | 1.85 | 1.88 | 147 |
| 3 | 6.2-7.3 | 3.52 | 3.57 | 3.24 | 3.29 | 128 |
| 4 | 7.4-15.0 | 8.04 | 10.41 | 6.90 | 9.02 | 361 |

**Figure 3.**
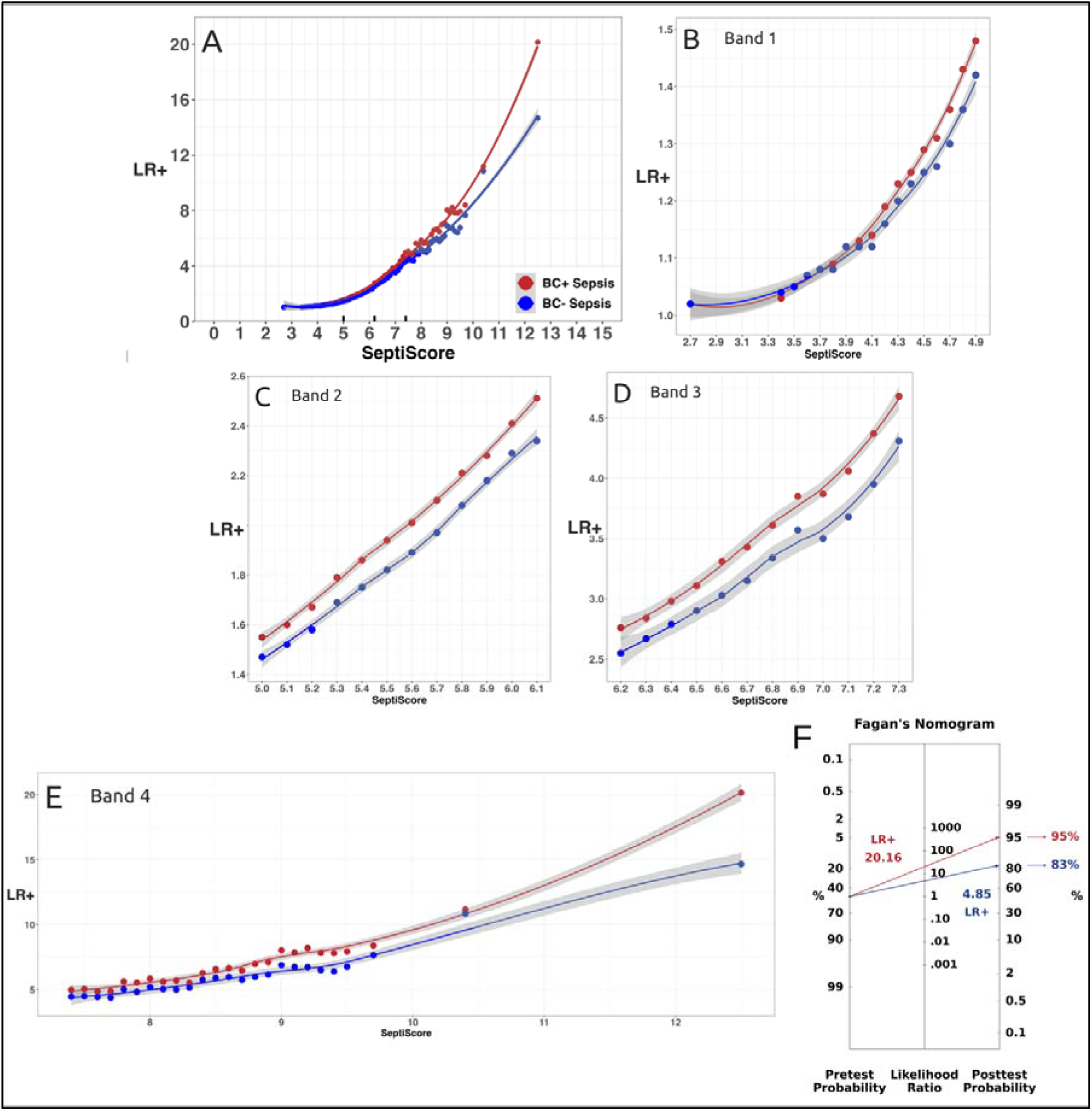
LR+ analysis for BC+ sepsis vs SIRS, and for BC-sepsis vs SIRS. **(A)** LR+ for 184 BC(+) sepsis vs. 386 SIRS (red line), and LR+ for 219 BC(−) sepsis vs. 386 SIRS (blue line). **(B)** Expanded view of Band 1. **C)** Expanded view of Band 2. **(D)** Expanded view of Band 3. **(E)** Expanded view of Band 4. For panels A-E the LOESS best fit lines and 95% error envelopes are shown. **(F)** Fagan nomogram for SeptiScore Band 4, assuming a pre-test sepsis prevalence of 50% and LR+ values of 4.85 (blue line) and 20.16 (red line). (These LR+ values correspond to the Band 4 lower and upper bounds, respectively.)

In Figure 3, **Panel A** shows the LR+ vs. SeptiScore relationship for 184 BC(+) sepsis vs 386 SIRS (red line), and also the analogous relationship for 219 BC(−) sepsis vs 386 SIRS (blue line). In both cases the LR+ increases monotonically with increasing SeptiScore, indicating that elevated SeptiScores are associated with greater likelihoods of sepsis. **Panels B, C, D and E** show expansions of the regions within Bands 1, 2, 3, 4 respectively. It is evident from the close-ups shown in these panels that the BC(+)sepsis vs. SIRS curve falls above the BC(−)sepsis vs. SIRS curve for all SeptiScores ≥ 4.0. **Panel F** shows a Fagan nomogram for Band 4 BC(+) sepsis vs. SIRS, assuming pre-test probability of 50% and using the minimum (4.85) and maximum (20.16) LR(+) values across this Band.

### 3.3. Negative Likelihood Ratios (LR-)

**Table 4** and **Figure 4** explore, for the entire available sample set of sepsis and SIRS/no sepsis patients (4 cohorts, N=889) the relationship between the SeptiScore and the negative likelihood ratio (LR-). No stratification according to BC(+/−) was performed. **Table 4** summarizes the LR-values at the lower and upper boundaries of each band, and also the mean and median LR-values for each band.

**Table 4.** LR-analysis for sepsis/SIRS discrimination, per SeptiScore Band. The dataset consists of 889 patient samples, without stratification by BC(+/−) status. The minimum, maximum, mean and median LR-values for each SeptiScore Band were estimated from the distributions provided in Supplement S1.

| Band | Range | LR- min, or at<br>lower boundary | median LR- | mean LR- | LR- max, or at<br>upper boundary | N |
| --- | --- | --- | --- | --- | --- | --- |
| 1 | 0.0-4.9 | 0.014 | 0.085 | 0.073 | 0.158 | 164 |
| 2 | 5.0-6.1 | 0.136 | 0.16 | 0.16 | 0.202 | 159 |
| 3 | 6.2-7.3 | 0.201 | 0.25 | 0.26 | 0.344 | 150 |
| 4 | 7.4-15.0 | 0.349 | 0.81 | 0.76 | 0.944 | 416 |

**Figure 4.**
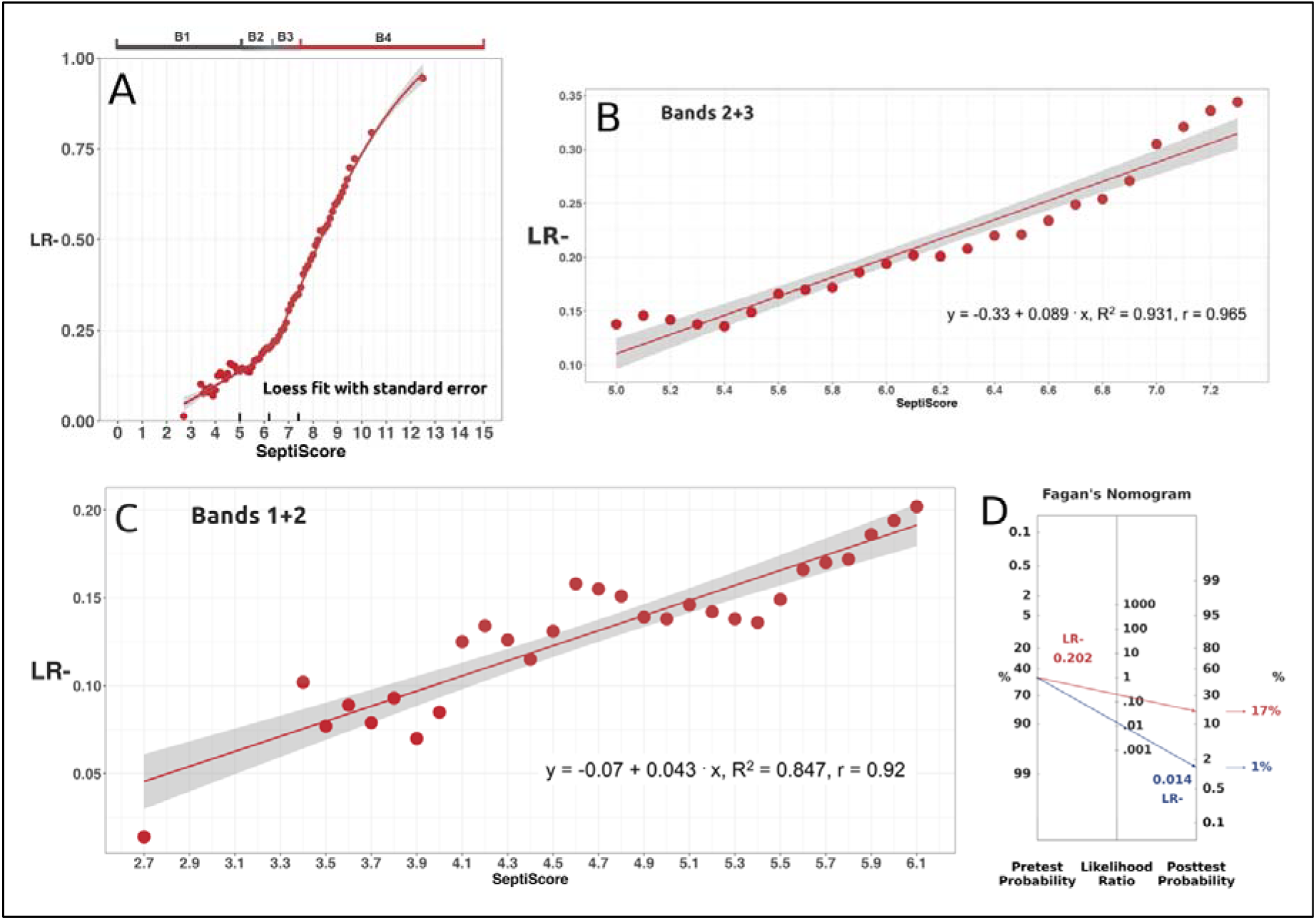
Negative likelihood ratio analysis. **(A)** LR-for all sepsis vs SIRS (4 cohorts, N=889) **(B)** Expanded region of LR-plot for SeptiScore Bands 2+3. **(C)** Expanded region of LR-plot for SeptiScore Bands 1+2. **(D)** Fagan’s nomogram for LR-SeptiScore Bands 1+2, showing the shift in sepsis probability from pre-test (p 0.50) to post-test (p 0.17 or 0.01) for LR-0.202 (red line) and 0.014 (blue line) respectively.

Figure 4. explores the relationship between SeptiScore and the negative likelihood ratio (LR-) across the SeptiScore range. All septic patients were included: there was no stratification according to whether the septic patients were BC(+), BC(−), or BC unknown. As shown in **Panel A** for a total of 503 sepsis vs. 386 SIRS patients, the LR-decreases monotonically with decreasing SeptiScore. Low SeptiScores are associated with markedly lower likelihoods of sepsis. **Panel B** shows an expanded region of the LR-versus SeptiScore plot for SeptiScore Bands 2+3. **Panel C** shows an expanded region of the LR-versus SeptiScore plot for SeptiScore Bands 1+2. **Panel D** shows Fagan’s LR-nomogram for SeptiScore Bands 1+2, assuming pre-test sepsis probability 50% and using the low (0.014, blue line) and high (0.202, red line) LR-values across these bands.

**Table 5** and **Figure 5** stratify the above analysis according to whether the septic patients can be classified as BC(+) or BC(−). (A total of 100 septic patients without reported BC data have been excluded in this analysis.)

**Table 5.** LR-analysis per SeptiScore Band, stratified according to whether BC(+) sepsis or BC(−) sepsis is being compared to SIRS. For this analysis, 789 patient samples were used, consisting of 184 Sepsis BC(+), 219 Sepsis BC(−) and 386 SIRS.

| SeptiScore<br>Band | SeptiScore<br>Range | LR- for<br>BC+ sepsis vs SIRS |  |  | LR- for<br>BC- sepsis vs SIRS |  |  |
| --- | --- | --- | --- | --- | --- | --- | --- |
|  |  | median | mean | N | median | mean | N |
| 1 | 0.0-4.9 | 0.022 | 0.030 | 140 | 0.107 | 0.09 | 152 |
| 2 | 5.0-6.1 | 0.043 | 0.055 | 129 | 0.167 | 0.168 | 135 |
| 3 | 6.2-7.3 | 0.166 | 0.175 | 95 | 0.258 | 0.266 | 102 |
| 4 | 7.4-15.0 | 0.745 | 0.695 | 206 | 0.755 | 0.730 | 216 |
|  |  | total |  | 570 |  |  | 605 |

**Figure 5.**
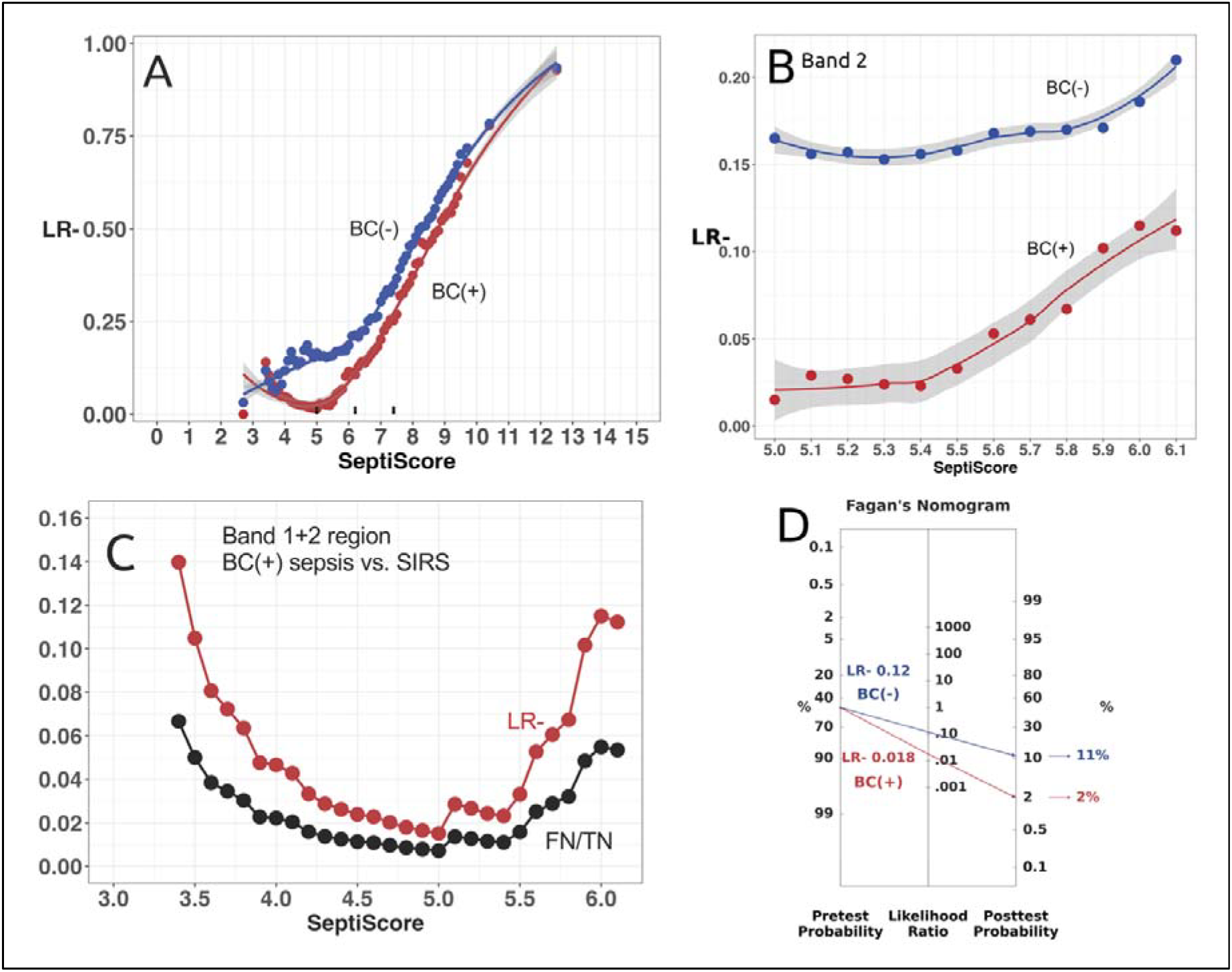
Negative likelihood ratio (LR-) stratified according to BC+ or BC-status. **(A)** Red: LR-for BC(+) sepsis (N=184) vs. SIRS (N=386). Blue: LR-for BC(−) sepsis (N=219) sepsis vs. SIRS (N=386). **(B)** Expanded Band 2 region of LR-plot. **(C)** BC(+) sepsis vs. SIRS comparison, expanded Band 1-2 region, showing the “J-shape” behavior (see Supplement S5). **(D)** Fagan’s nomogram at LR-minima, assuming pre-test sepsis probability 50%. For BC(−), the minimum value LR− = 0.12 occurs at SeptiScore 3.4 (blue line in panel A). For BC(+), the minimum value LR− = 0.018 occurs at SeptiScore 5.0 (red line in panel A).

Figure 5. explores further the relationship between SeptiScore and the negative likelihood ratio (LR-), after stratifying by whether BC(+) sepsis or BC(−) sepsis is being compared to SIRS. Note that in BC(+) case we observe a distinctive *J-shaped* curve with a minimum near 5.0 and an increasing trend as the SeptiScore decreases from 5.0 to 3.4 (red points in panels 5A, 5C). This trend is mirrored by the FN/TN ratio (black points in panel 5C) and is readily explained as a consequence of small sample size in the Band 1 SeptiScore region (see **Supplement S5**). Consequently, due to the paucity of sepsis BC(+) cases in Band 1, LR-estimates in Band 1 are not very precise.

## 4. Discussion

This study has addressed the previously stated four hypotheses. Our analyses show that the likelihood ratio (LR) framework can provide a clinically useful interpretive approach that complements the previously used SeptiScore banding scheme. Low Band 1 SeptiScores are associated with sufficiently small LR-to support the use of SeptiCyte RAPID as a rule-out test for sepsis. Band 2 SeptiScores have a median LR-of 0.16 (range 0.14-0.20) indicating a moderate decrease in the likelihood of sepsis. High Band 4 SeptiScores are associated with sufficiently large LR+ to support the use of SeptiCyte RAPID as a rule-in test for sepsis. Band 3 SeptiScores have a median LR+ of 3.26 (range 2.57-4.24) indicating an increase in the likelihood of sepsis per the interpretation of LR+ in Elkahwagy *et al* (2024) [12]. SeptiScore-derived LR+ and LR-values can be combined with estimates of pre-test probability (derived from patient characteristics and/or other diagnostic tests) to generate individualized, patient-specific post-test probabilities of sepsis.

Applying likelihood ratio (LR) analysis to SeptiCyte RAPID enhances the clinical interpretation of the SeptiScore for discriminating sepsis from SIRS. Unlike simple sensitivity and specificity metrics, LRs provide a direct measure of how a specific test result modifies the clinician’s *a prioi* estimated probability of disease in an individual patient. Positive likelihood ratios (LR+) quantify how much a high SeptiScore (in Band 3 or 4) increases the likelihood of sepsis, while negative likelihood ratios (LR-) indicate how much a low SeptiScore (in Band 1 or 2) decreases it. Of course, SeptiCyte RAPID should be considered an *aid* to diagnosing sepsis, and therefore SeptiScores must be taken into consideration alongside other laboratory test results and vital signs.

### 4.1 Clinical Implications of High LR+ Values

High positive likelihood ratios (LR+) for band 4 SeptiScores, which precede traditional microbiology results, may provide clinicians with early confidence of a sepsis diagnosis, and may support microbiology diagnostic stewardship.

Band 3 SeptiScores had a median LR+ 3.26 while Band 4 SeptiScores had a median LR+ 6.97. At the upper end of Band 4, the LR+ had a value of 15.57 (Fig 2E). At SeptiScores ≥ 11.4, the LR+ ≥10 for the sepsis/SIRS discrimination. These results provide strong evidence to rule in sepsis at high SeptiScores.

When stratified on blood culture results, some differences in LR+ were evident between the BC+ sepsis/SIRS and BC-sepsis/SIRS comparisons. For the BC+ case, Band 3 and 4 SeptiScores had median LR+ of 3.52 and 8.04 respectively, while for the BC-case, the corresponding values were 3.24 and 6.90. For SeptiScores ≥ 11.4, the LR+ ≥10 for BC+ sepsis. High SeptiScore values therefore may suggest that a blood culture be drawn prior to antibiotics being started.

### 4.2. Clinical Implications of Low LR-Values

For sepsis/SIRS discrimination in Band 1, the median LR-was 0.014 irrespective of BC status, 0.022 for BC+ sepsis and 0.107 for BC-sepsis respectively. These results indicate in general a very low probability of sepsis associated with Band 1 SeptiScores. Low LR-values for Band 1 SeptiScores may prompt clinicians to consider alternate diagnoses to sepsis, and might also lead to better antibiotic stewardship. Such results, obtained early in the diagnostic workup process, may lead to fewer missed diagnoses and more efficient use of hospital resources.

### 4.3. Comparing SeptiCyte RAPID to Other Blood-Based Immune Response Sepsis Biomarkers

To provide further context, **Tables 6, 7** summarize LR+ and LR-data from the literature for other blood-based immune response biomarkers that have been used as aids in sepsis diagnosis. These comparative tables demonstrate that the performance of SeptiCyte RAPID is relatively strong at both the high end (LR+) and the low end (LR-) of the 0-15 SeptiScore range.

**Table 6.**
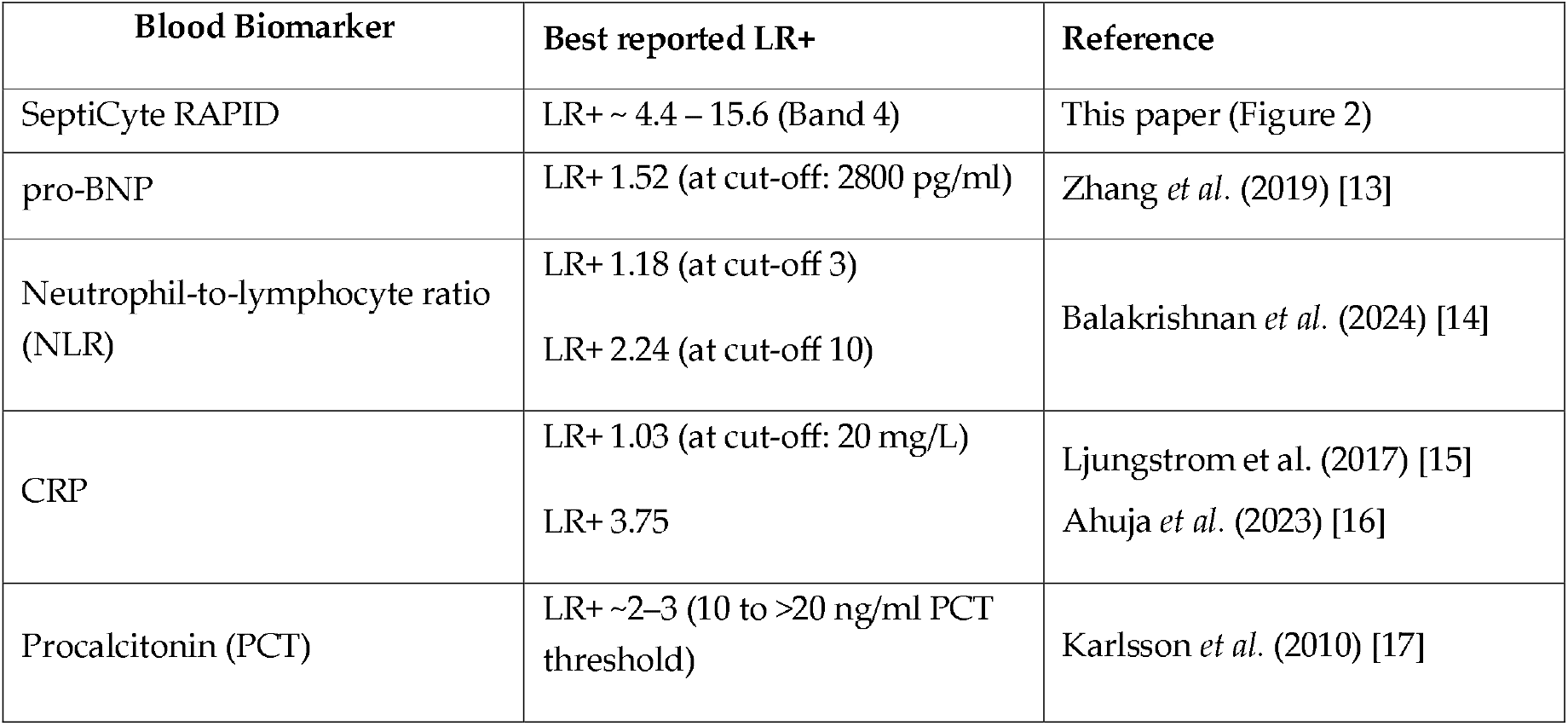
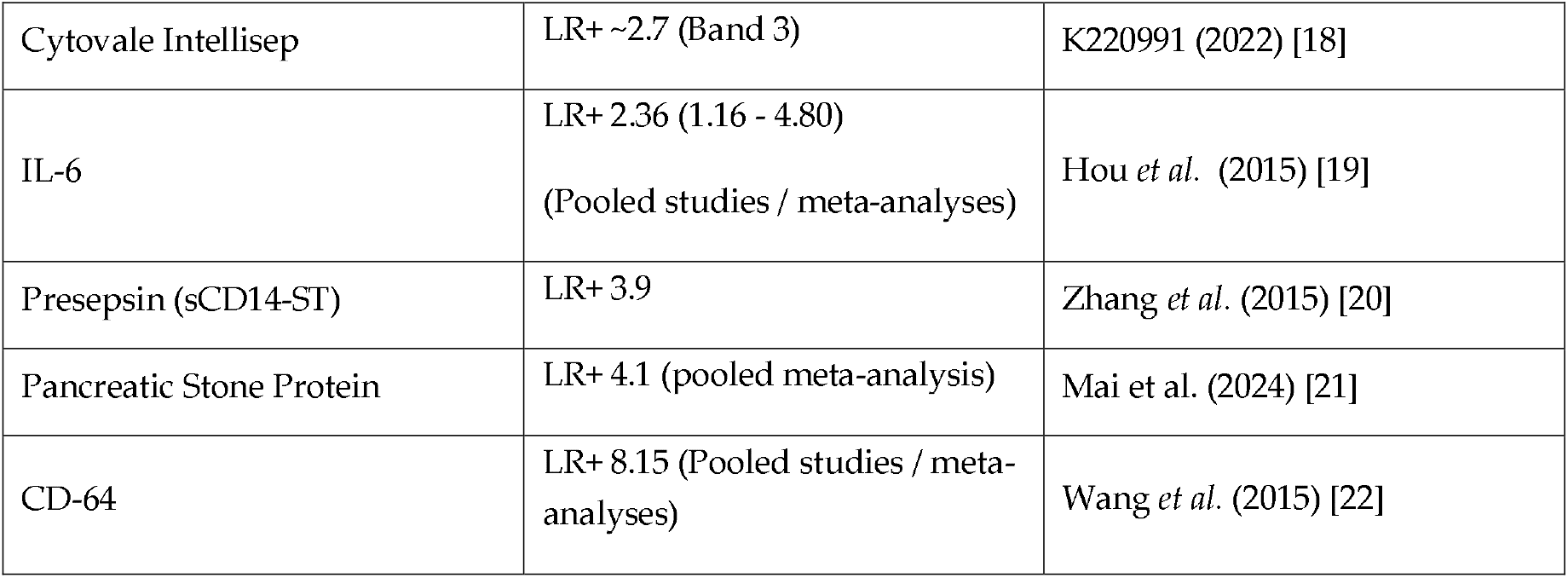
Comparative LR+ ranges for blood-based immune response sepsis biomarkers. Data are summarized from the present paper (SeptiCyte RAPID), from FDA Decision Summary (Cytovale Intellisep), or the scientific literature (others).

**Table 7.**
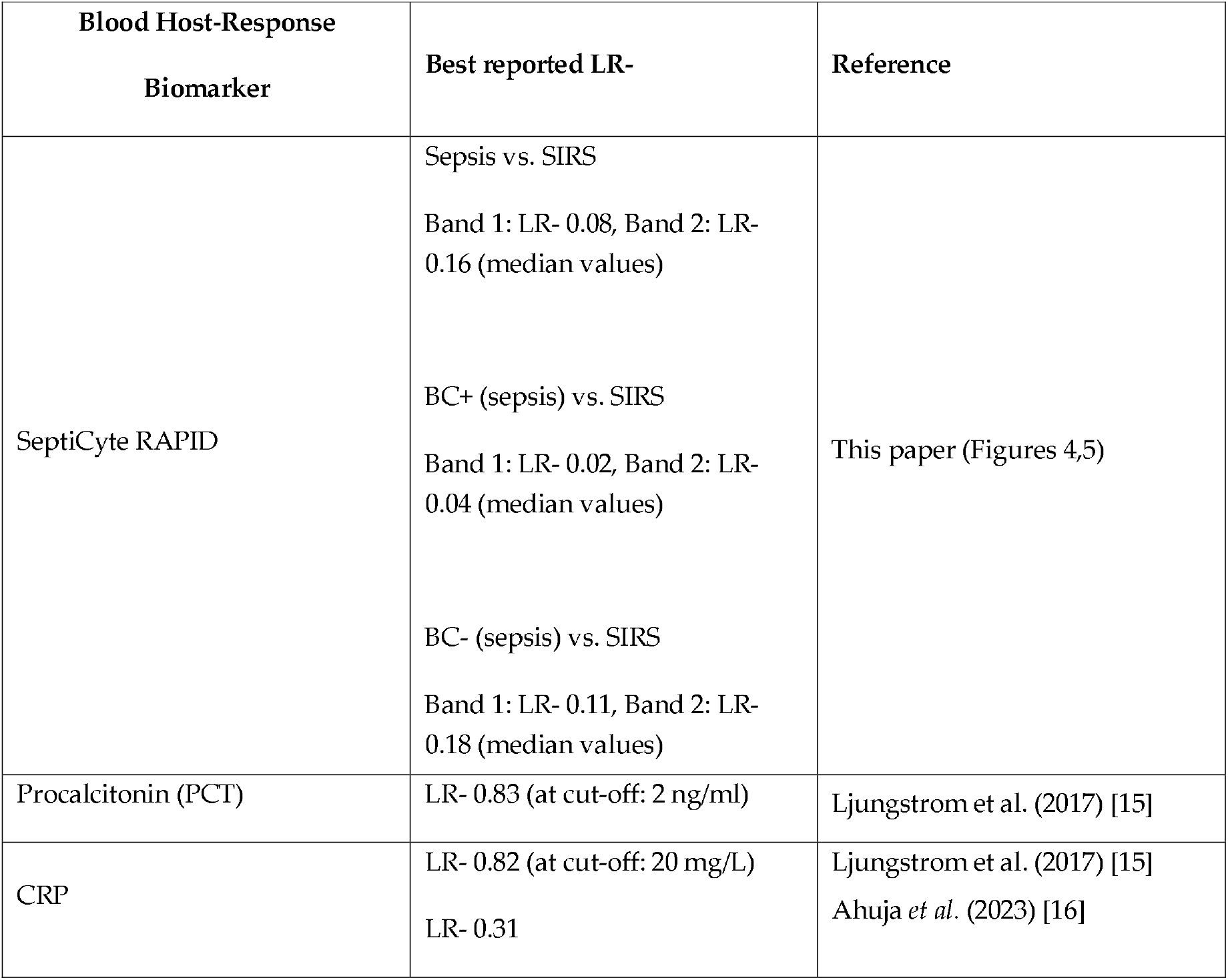
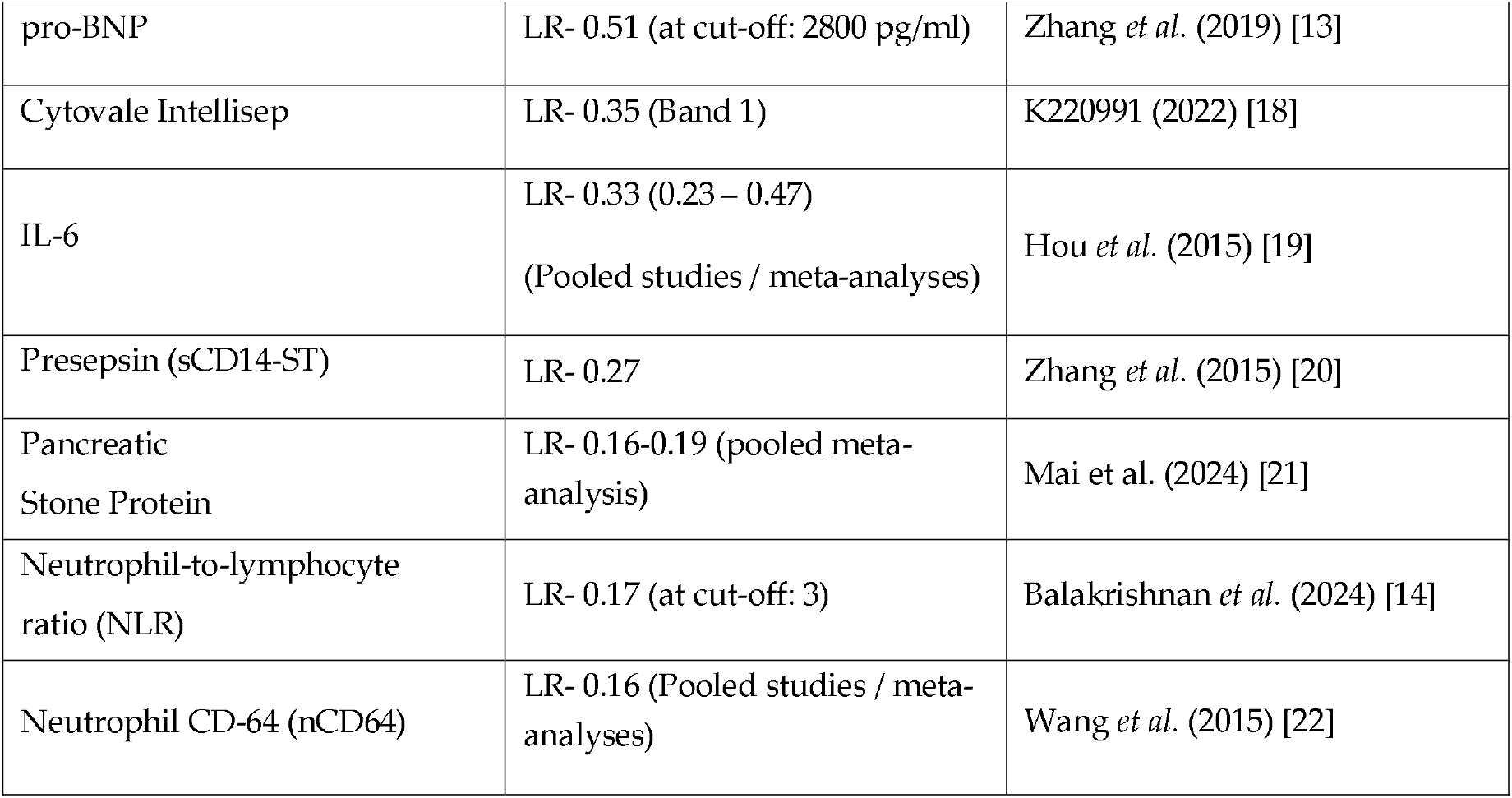
Comparative LR-ranges for blood-based immune response sepsis biomarkers. Data are summarized from the present paper (SeptiCyte RAPID), from FDA Decision Summary (Cytovale Intellisep), or the scientific literature (others).

| Blood Host-Response<br>Biomarker | Best reported LR- | Reference |
| --- | --- | --- |
| SeptiCyt RAPID | Sepsis vs. SIRS<br>Band 1: LR- 0.08, Band 2: LR- 0.16 (median values)<br>BC+ (sepsis) vs. SIRS<br>Band 1: LR- 0.02, Band 2: LR- 0.04 (median values)<br>BC- (sepsis) vs. SIRS<br>Band 1: LR- 0.11, Band 2: LR- 0.18 (median values) | This paper (Figures 4,5) |
| Procalcitonin (PCT) | LR- 0.83 (at cut-off: 2 ng/ml) | Ljungstrom *et al.* (2017) [15] |
| CRP | LR- 0.82 (at cut-off: 20 mg/L)<br>LR- 0.31 | Ljungstrom *et al.* (2017) [15]<br>Ahuja *et al.* (2023) [16] |
| pro-BNP | LR- 0.51 (at cut-off: 2800 pg/ml) | Zhang *et al.* (2019) [13] |
| Cytovale Intellisep | LR- 0.35 (Band 1) | K220991 (2022) [18] |
| IL-6 | LR- 0.33 (0.23 – 0.47)<br>(Pooled studies / meta-analyses) | Hou *et al.* (2015) [19] |
| Presepsin (sCD14-ST) | LR- 0.27 | Zhang *et al.* (2015) [20] |
| Pancreatic<br>Stone Protein | LR- 0.16-0.19 (pooled meta-<br>analysis) | Mai *et al.* (2024) [21] |
| Neutrophil-to-lymphocyte<br>ratio (NLR) | LR- 0.17 (at cut-off: 3) | Balakrishnan *et al.* (2024) [14] |
| Neutrophil CD-64 (nCD64) | LR- 0.16 (Pooled studies / meta-<br>analyses) | Wang *et al.* (2015) [22] |

### 4.4. Applying Likelihood Ratio Analysis to Individual Patients

In applying likelihood ratio–based analysis of sepsis versus SIRS to individual patients, a key parameter is the pre-test probability, which must be specified *a priori* and directly influences the resulting post-test probability estimates. In diagnostic terms, this represents the estimated likelihood that a patient has sepsis at the time of initial clinical assessment, prior to incorporating additional test results.

Our analysis estimates sepsis likelihood at the cohort level, using aggregated data to derive both pre-test probabilities and likelihood ratios (LRs). In clinical practice, however, clinicians typically assess sepsis risk on an individual basis, incorporating patient-specific factors such as age, comorbidities, vital signs, laboratory findings, and other contextual information. For likelihood ratio–based (Bayesian) approaches to be clinically informative, the pre-test probability should therefore be individualized rather than population-based.

In the absence of detailed patient-level data, the pre-test probability may be approximated by the point prevalence of sepsis in the source population. Accordingly, we assumed an average prevalence of 50% ± 2.5% across all clinical sites in our study (Supplement S6). This estimate exceeds the range reported in the ICON audit [23], suggesting the possibility of selection bias in our cohort; this limitation is discussed further below.

Recent studies have demonstrated approaches to estimating patient-specific sepsis risk using multivariable prediction models. For example, nomogram-based models integrating clinical features and comorbidities have been developed for general ICU populations [24], for patients using routinely collected vital signs and laboratory data [25], for urinary tract infection–associated sepsis [26], and for community-acquired pneumonia [27].

These tools illustrate how individualized risk estimation can provide a practical foundation for integrating likelihood ratio–based reasoning into clinical decision-making. Such models could be extended to incorporate SeptiCyte RAPID scores, enabling a combined approach to estimating sepsis probability. Development of this integrated framework is the subject of ongoing work.

### 4.5. Limitations

We have identified some limitations to this study. First, the analysis is restricted to ICU patients. Although the cohort includes patients from 19 hospitals, the generalizability of these findings to patients presenting in the emergency department or hospital wards has not been established. In addition, the racial and ethnic composition of the cohort is uneven, with overrepresentation of White patients (67%) and relatively low representation of Hispanic and Asian patients (3.5–3.6% each). The available sample size does not support reliable stratified analyses by race or ethnicity.

Second, there is likely variability in the accuracy of the reference standard (sepsis vs. SIRS classification) across participating sites. Adjudications were based on a combination of Sepsis-2 criteria and Sepsis-3 criteria frameworks, which may introduce heterogeneity in diagnostic classification. Nevertheless, all cases were reviewed by qualified clinicians, who applied locally accepted criteria based on expert judgment.

Third, the group of patients classified as SIRS is clinically heterogeneous. Although all met the inclusion criterion of at least two SIRS features, patients varied widely in age, comorbidities, and other clinical characteristics. This heterogeneity may introduce additional variability into the analysis; however, it also provides a degree of robustness, as likelihood ratio–based performance appears to be maintained across a diverse patient population.

Fourth, estimates of the pre-test probability of sepsis (operationalized here as cohort-specific prevalence) vary substantially across our cohorts. Overall sepsis prevalence is approximately 42% in the 510k cohort, compared with approximately 75% in the Andalusian cohort. This heterogeneity likely reflects a combination of selection bias, differences in sepsis versus SIRS adjudication criteria, and variation in baseline illness severity across component sites within our cohorts.

Fifth, relatively few patients are represented at the extremes of the SeptiScore distribution. As a result, likelihood ratios (LRs) in these regions were estimated using interval-based approaches rather than point estimates, leading to reduced resolution and wider confidence intervals at the tails of the LR versus SeptiScore relationship.

## 5. Conclusions

The application of a likelihood ratio framework to SeptiCyte RAPID results provides a clinically actionable method for differentiating sepsis from SIRS in critically ill patients. By integrating these ratios with patient-specific pre-test probabilities, clinicians can more accurately quantify the shift in disease likelihood following the test.

## Data Availability

All data produced in the present study are available upon reasonable request to the authors

## Supplementary Materials

S1: Definition of SeptiScore band boundaries. S2: Numbers of SIRS & sepsis patients per SeptiScore interval, in the LR analysis of sepsis vs. SIRS. S3: Numbers of SIRS & sepsis patients per SeptiScore interval, in the LR analysis of BC(+) sepsis vs. SIRS. S4: Numbers of SIRS & sepsis patients per SeptiScore interval, in the LR analysis of BC(−) sepsis vs. SIRS. S5: Further analysis of J-shaped curve in Figure 5C. S6: Comparative estimates of sepsis pre-test probability.

## Author Contributions

**Conceptualization**, Krupa Navalkar, Thomas Yager, Richard Brandon and Prashant Wani; **Data curation**, Thomas Yager and Silvia Cermelli; **Formal analysis**, Krupa Navalkar, Thomas Yager and Richard Brandon; **Funding acquisition**, José Garnacho-Montero, María Cantón-Bulnes, Maximilian Dietrich, Maik von der Forst, Sören L. Becker, Sophia Benthien; **Investigation**, Krupa Navalkar and Thomas Yager; **Methodology**, Krupa Navalkar, Thomas Yager, Roy Davis, Prashant Wani and Richard Brandon; **Project administration**, José Garnacho-Montero, María Cantón-Bulnes, Richard Rothman, Russell R. Miller III, John Burke, Maximilian Dietrich, Maik von der Forst, Sören L. Becker, Sophia Benthien and Richard Brandon; **Software**, Krupa Navalkar; **Supervision**, José Garnacho-Montero, María Cantón-Bulnes, Maximilian Dietrich, Maik von der Forst, Sören L. Becker, Sophia Benthien, Thomas Yager, Roy Davis, Silvia Cermelli and Richard Brandon; **Validation**, José Garnacho-Montero, María Luisa Cantón-Bulnes, José Luis García-Garmendia, Ángel Estella, Adela Fernández-Galilea, Russell R. Miller III, John Burke, Gourang Patel, Jorge Parada, Marcus J. Schultz, Maximilian Dietrich, Maik von der Forst, Sören L. Becker, Sophia Benthien, Elisa Baumann, Carsten Zeiner, Philipp M. Lepper, Richard Rothman; **Visualization**, Krupa Navalkar; **Writing – original draft**, Thomas Yager and Richard Brandon; **Writing – review & editing**, Krupa Navalkar, Gourang Patel, Jorge Parada, Marcus J. Schultz, Thomas Yager, Prashant Wani, Roy Davis, Silvia Cermelli and Richard Brandon.

## Institutional Review Board Statement

The studies were conducted in accordance with the Declaration of Helsinki and approved by Institutional Review Boards (see below).

The 510K cohort was drawn from retrospective (NCT01905033 and NCT02127502; clinicaltrials.gov (accessed on 17 February 2024)) and prospective trials (NCT05469048; clinicaltrials.gov (first submitted 15 July 2022, accessed on 17 February 2024)). The studies were called MARS, VENUS and NEPTUNE. Ethics approval for the MARS trial was given by the Medical Ethics Committee of the Amsterdam Medical Center (approval 156 June 2010, # 10-056C). Ethics approvals for the VENUS trial were given by the relevant Institutional Review Boards as follows: Intermountain Medical Center/Latter Day Saints Hospital (approval, 21 February 2016, # 1024931); Johns Hopkins Hospital (approval 28 January 2016, # IRB00087839); Rush University Medical Center (approval 11 March 2016, # 15111104-IRB01); Loyola University Medical Center (approval, 10 March 2016, # 208291); Northwell Healthcare (approval 1 April 2016, #16-02-42-03). Ethics approvals for the NEPTUNE trial were given by the relevant Institutional Review Boards as follows: Emory University (approval 4 December 2019, # IRB00115400); Grady Memorial Hospital (approval, 14 January 2020, # 00-115400); Rush University Medical Center (approval, 16 January 2020, # 19101603-IRB01); University of Southern California Medical Center (approval, 10 February 2020, # HS-19-0884-CR001). The Andalusian study was conducted in seven ICUs in Spain and coordinated by the Virgen Macarena University Hospital in Seville. The study was approved by the Research Ethics Committees of the Virgen Macarena-Virgen Rocio University Hospitals (Certificate of Ethics Approval dated 23 Dec 2021. Trial name SEPT-ANC. Codigo Interno: 2662-N-21.) The SeptAsTERS component of this study was approved by the Ethics Committee of the Medical Faculty of Heidelberg University (S-118/2021; approval date 4 January 2021) and was registered in the German Clinical Trials Register (DRKS00024891; registration date 4 August 2021) prior to enrolment. The study was approved by the Ethics Committee of the Saarland Medical Association for Saarland University (approval # 68/22, date 23 May 2022).

## Informed Consent Statement

Informed consent was obtained from all subjects involved in the clinical studies. All methods used in this study were carried out in accordance with the relevant guidelines and regulations. For the secondary cohort blood sample extractions were permitted prior to obtaining written permission, if needed, to enable SeptiCyte RAPID results to be generated as quickly as possible. Written consent from the patient, or next of kin, was obtained within 48 hours of ICU admission and the blood sample was discarded if written consent was not obtained.

## Data Availability Statement

The datasets presented in this article are not readily available for patient privacy and commercial reasons. Requests to access the datasets should be directed to Richard Brandon.

