## Supplementary File for "Ruling In and Ruling Out Sepsis Using Likelihood Ratios of a Host Response Assay"

**SUPPLEMENTARY MATERIAL**

| **Supplement #** | **Topic** |
|---|---|
| S1 | Description of SeptiScore Band Boundaries |
| S2 | Numbers of SIRS & sepsis patients per SeptiScore interval, in the LR analysis of sepsis vs. SIRS |
| S3 | Numbers of SIRS & sepsis patients per SeptiScore interval, in the LR analysis of BC(+) sepsis vs. SIRS |
| S4 | Numbers of SIRS & sepsis patients per SeptiScore interval, in the LR analysis of BC(-) sepsis vs. SIRS |
| S5 | Further analysis of J-shaped Curve in Figure 5C |
| S6 | Comparative estimates of pre-test probability of sepsis |

**Supplement S1: Description of SeptiScore Band Boundaries**

Much of the analysis and discussion in this paper centers around the concept of "SeptiScore Bands". Four SeptiScore Bands and their boundaries had been pre-defined, in the following way, as described previously in Balk et al. (2024) [1]. An independent set of 195 patient samples that had been adjudicated by RPD was used to set the Band 1/2 cutoff at 5.0 to achieve 90% sensitivity, and the Band 3/4 cutoff at 7.4 to achieve 80% specificity. The Band 2/3 cutoff was then defined as the midpoint between these two outer thresholds.

For descriptive comparison, Figure S1 shows the relationship between LR+ and SeptiScore on a semilogarithmic scale. This visualization indicates that the previously defined Band 1/2 and Band 3/4 boundaries fall in regions where the slope of the log(LR+) versus SeptiScore curve appears to change. This observation is presented as a qualitative consistency check between the pre-specified band structure and the behavior of LR+ across the SeptiScore range.

**Figure S1: Plot of log(LR+) as a function of SeptiScore**

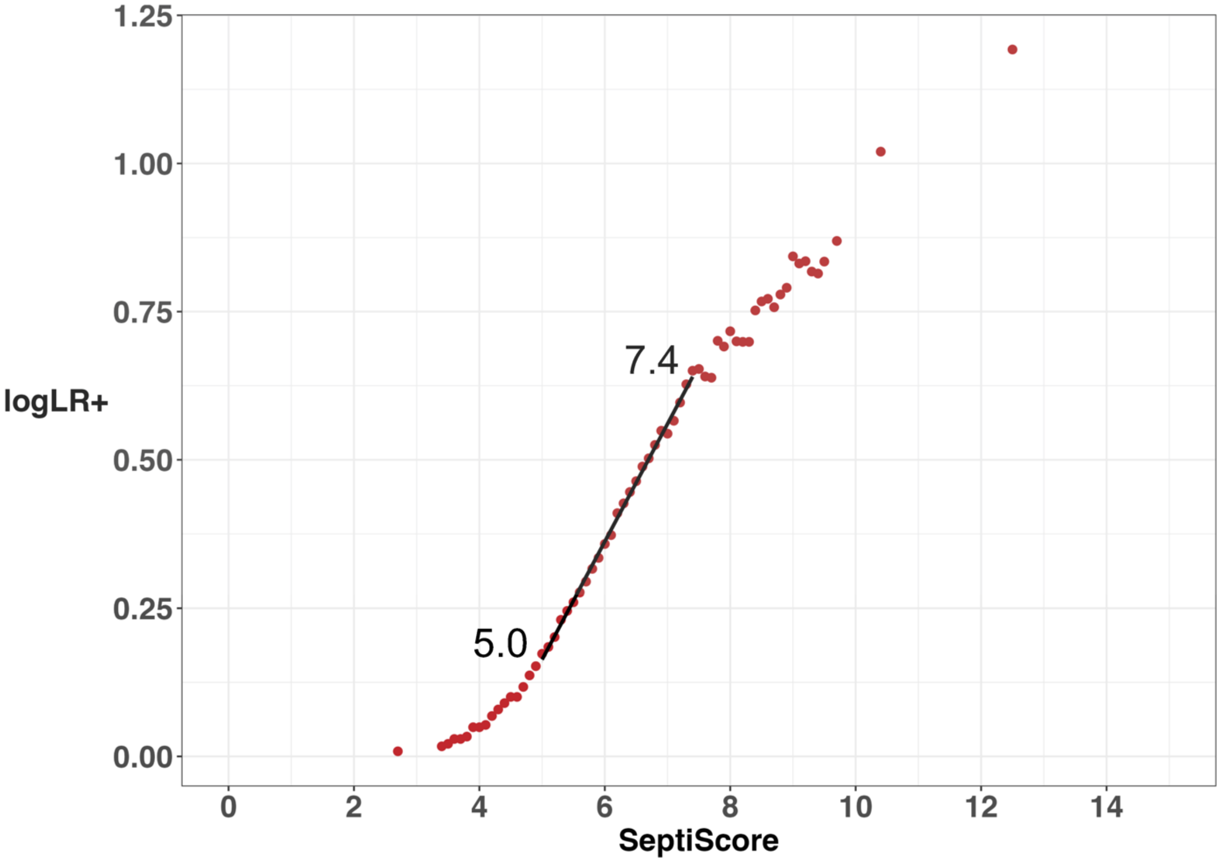


**Supplement S2:** Numbers of sepsis & SIRS patients per SeptiScore interval, in the LR analysis of sepsis vs. SIRS. The dataset for this analysis consists of N=889 patients. Likelihood ratio estimates at the extreme ends of the SeptiScore distribution are based on wide SeptiScore intervals to allow sufficient numbers of SIRS & sepsis patients per interval for accurate LR estimation.

| SeptiScore range (midpoint) | RAPID Interpretation Band | Likelihood ratio (positive) | Likelihood ratio (negative) | No. of Sepsis patients | No. of SIRS patients | Total No. patients |
|---|---|---|---|---|---|---|
| 1.8-3.3 (mid 2.7) | Band 1 | 1.02 | 0.014 | 2 | 15 | 17 |
| 3.4 | Band 1 | 1.04 | 0.102 | 0 | 5 | 5 |
| 3.5 | Band 1 | 1.05 | 0.077 | 1 | 6 | 7 |
| 3.6 | Band 1 | 1.07 | 0.089 | 0 | 3 | 3 |
| 3.7 | Band 1 | 1.07 | 0.079 | 1 | 4 | 5 |
| 3.8 | Band 1 | 1.08 | 0.093 | 0 | 11 | 11 |
| 3.9 | Band 1 | 1.12 | 0.070 | 1 | 1 | 2 |
| 4 | Band 1 | 1.12 | 0.085 | 3 | 4 | 7 |

| SeptiScore range (midpoint) | RAPID Interpretation Band | Likelihood ratio (positive) | Likelihood ratio (negative) | No. of Sepsis patients | No. of SIRS patients | Total No. patients |
|---|---|---|---|---|---|---|
| 4.1 | Band 1 | 1.13 | 0.125 | 3 | 14 | 17 |
| 4.2 | Band 1 | 1.17 | 0.134 | 1 | 10 | 11 |
| 4.3 | Band 1 | 1.2 | 0.126 | 0 | 7 | 7 |
| 4.4 | Band 1 | 1.23 | 0.115 | 3 | 8 | 11 |
| 4.5 | Band 1 | 1.26 | 0.131 | 4 | 4 | 8 |
| 4.6 | Band 1 | 1.26 | 0.158 | 2 | 12 | 14 |
| 4.7 | Band 1 | 1.31 | 0.155 | 2 | 13 | 15 |
| 4.8 | Band 1 | 1.37 | 0.151 | 0 | 10 | 10 |
| 4.9 | Band 1 | 1.42 | 0.139 | 2 | 12 | 14 |
| 5 | Band 2 | 1.49 | 0.138 | 3 | 8 | 11 |
| 5.1 | Band 2 | 1.53 | 0.146 | 1 | 10 | 11 |
| 5.2 | Band 2 | 1.59 | 0.142 | 2 | 16 | 18 |
| 5.3 | Band 2 | 1.7 | 0.138 | 1 | 8 | 9 |
| 5.4 | Band 2 | 1.76 | 0.136 | 5 | 9 | 14 |
| 5.5 | Band 2 | 1.82 | 0.149 | 6 | 9 | 15 |
| 5.6 | Band 2 | 1.89 | 0.166 | 3 | 9 | 12 |
| 5.7 | Band 2 | 1.97 | 0.170 | 3 | 10 | 13 |
| 5.8 | Band 2 | 2.07 | 0.172 | 6 | 9 | 15 |
| 5.9 | Band 2 | 2.16 | 0.186 | 5 | 10 | 15 |
| 6 | Band 2 | 2.28 | 0.194 | 4 | 6 | 10 |
| 6.1 | Band 2 | 2.36 | 0.202 | 3 | 13 | 16 |
| 6.2 | Band 3 | 2.57 | 0.201 | 4 | 6 | 10 |
| 6.3 | Band 3 | 2.67 | 0.208 | 6 | 7 | 13 |
| 6.4 | Band 3 | 2.79 | 0.220 | 2 | 5 | 7 |
| 6.5 | Band 3 | 2.91 | 0.221 | 7 | 8 | 15 |
| 6.6 | Band 3 | 3.08 | 0.234 | 7 | 5 | 12 |
| 6.7 | Band 3 | 3.18 | 0.249 | 4 | 6 | 10 |
| 6.8 | Band 3 | 3.35 | 0.254 | 9 | 7 | 16 |
| 6.9 | Band 3 | 3.54 | 0.271 | 14 | 2 | 16 |
| 7 | Band 3 | 3.5 | 0.305 | 9 | 6 | 15 |
| 7.1 | Band 3 | 3.68 | 0.321 | 9 | 7 | 16 |
| 7.2 | Band 3 | 3.95 | 0.336 | 6 | 6 | 12 |
| 7.3 | Band 3 | 4.24 | 0.344 | 4 | 4 | 8 |
| 7.4 | Band 4 | 4.47 | 0.349 | 9 | 2 | 11 |
| 7.5 | Band 4 | 4.5 | 0.368 | 16 | 1 | 17 |
| 7.6 | Band 4 | 4.37 | 0.405 | 7 | 1 | 8 |

| SeptiScore range (midpoint) | RAPID Interpretation Band | Likelihood ratio (positive) | Likelihood ratio (negative) | No. of Sepsis patients | No. of SIRS patients | Total No. patients |
|---|---|---|---|---|---|---|
| 7.7 | Band 4 | 4.35 | 0.420 | 9 | 9 | 18 |
| 7.8 | Band 4 | 5.02 | 0.429 | 7 | 0 | 7 |
| 7.9 | Band 4 | 4.91 | 0.445 | 8 | 4 | 12 |
| 8 | Band 4 | 5.21 | 0.458 | 12 | 0 | 12 |
| 8.1 | Band 4 | 5.01 | 0.485 | 7 | 1 | 8 |
| 8.2 | Band 4 | 5 | 0.499 | 13 | 2 | 15 |
| 8.3 | Band 4 | 5 | 0.525 | 2 | 5 | 7 |
| 8.4 | Band 4 | 5.65 | 0.522 | 6 | 2 | 8 |
| 8.5 | Band 4 | 5.85 | 0.532 | 5 | 1 | 6 |
| 8.6 | Band 4 | 5.91 | 0.541 | 8 | 0 | 8 |
| 8.7 | Band 4 | 5.72 | 0.559 | 11 | 3 | 14 |
| 8.8 | Band 4 | 6.01 | 0.578 | 10 | 2 | 12 |
| 8.9 | Band 4 | 6.17 | 0.596 | 7 | 4 | 11 |
| 9 | Band 4 | 6.97 | 0.604 | 6 | 0 | 6 |
| 9.1 | Band 4 | 6.78 | 0.617 | 7 | 1 | 8 |
| 9.2 | Band 4 | 6.84 | 0.630 | 8 | 0 | 8 |
| 9.3 | Band 4 | 6.57 | 0.647 | 10 | 1 | 11 |
| 9.4 | Band 4 | 6.52 | 0.666 | 18 | 3 | 21 |
| 9.5 | Band 4 | 6.83 | 0.698 | 8 | 1 | 9 |
| 9.6-9.9 (mid 9.7) | Band 4 | 7.4 | 0.723 | 21 | 6 | 27 |
| 10.0-10.9 (mid 10.4) | Band 4 | 10.47 | 0.795 | 60 | 6 | 66 |
| 11.0-15.0 (mid 12.5) | Band 4 | 15.57 | 0.944 | 80 | 6 | 86 |
| | | | **Total** | **503** | **386** | **889** |

**Supplement S3:** Numbers of SIRS & sepsis patients per SeptiScore interval, in the LR analysis of BC(+) sepsis vs. SIRS. The dataset for this analysis consists of N=570 patients. Likelihood ratio estimates at the extreme ends of the SeptiScore distribution are based on wide SeptiScore intervals to allow sufficient numbers of SIRS & sepsis patients per interval for accurate LR estimation.

| SeptiScore range (midpoint) | RAPID Interpretation Band | Likelihood ratio (positive) | Likelihood ratio (negative) | No. of BC+ Sepsis patients | No. of SIRS patients | Total No. patients |
|---|---|---|---|---|---|---|
| 1.8-3.3 (mid 2.7) | Band 1 | 1.02 | 0.000 | 1 | 15 | 16 |
| 3.4 | Band 1 | 1.03 | 0.140 | 0 | 5 | 5 |
| 3.5 | Band 1 | 1.05 | 0.105 | 0 | 6 | 6 |
| 3.6 | Band 1 | 1.07 | 0.081 | 0 | 3 | 3 |
| 3.7 | Band 1 | 1.08 | 0.072 | 0 | 4 | 4 |
| 3.8 | Band 1 | 1.09 | 0.064 | 0 | 11 | 11 |
| 3.9 | Band 1 | 1.12 | 0.048 | 0 | 1 | 1 |
| 4 | Band 1 | 1.13 | 0.047 | 0 | 4 | 4 |
| 4.1 | Band 1 | 1.14 | 0.043 | 0 | 14 | 14 |
| 4.2 | Band 1 | 1.19 | 0.033 | 0 | 10 | 10 |
| 4.3 | Band 1 | 1.23 | 0.029 | 0 | 7 | 7 |
| 4.4 | Band 1 | 1.25 | 0.026 | 0 | 8 | 8 |
| 4.5 | Band 1 | 1.29 | 0.024 | 0 | 4 | 4 |
| 4.6 | Band 1 | 1.31 | 0.023 | 0 | 12 | 12 |
| 4.7 | Band 1 | 1.36 | 0.020 | 0 | 13 | 13 |
| 4.8 | Band 1 | 1.43 | 0.018 | 0 | 10 | 10 |
| 4.9 | Band 1 | 1.48 | 0.017 | 0 | 12 | 12 |
| 5 | Band 2 | 1.55 | 0.015 | 1 | 8 | 9 |
| 5.1 | Band 2 | 1.6 | 0.029 | 0 | 10 | 10 |
| 5.2 | Band 2 | 1.67 | 0.027 | 0 | 16 | 16 |
| 5.3 | Band 2 | 1.79 | 0.024 | 0 | 8 | 8 |
| 5.4 | Band 2 | 1.86 | 0.023 | 1 | 9 | 10 |
| 5.5 | Band 2 | 1.94 | 0.033 | 2 | 9 | 11 |
| 5.6 | Band 2 | 2.01 | 0.053 | 1 | 9 | 10 |
| 5.7 | Band 2 | 2.1 | 0.061 | 1 | 10 | 11 |
| 5.8 | Band 2 | 2.21 | 0.067 | 4 | 9 | 13 |
| 5.9 | Band 2 | 2.28 | 0.102 | 2 | 10 | 12 |
| 6 | Band 2 | 2.41 | 0.115 | 0 | 6 | 6 |
| 6.1 | Band 2 | 2.51 | 0.112 | 0 | 13 | 13 |
| 6.2 | Band 3 | 2.76 | 0.107 | 3 | 6 | 9 |
| 6.3 | Band 3 | 2.84 | 0.128 | 2 | 7 | 9 |
| 6.4 | Band 3 | 2.98 | 0.140 | 0 | 5 | 5 |
| 6.5 | Band 3 | 3.11 | 0.138 | 2 | 8 | 10 |
| 6.6 | Band 3 | 3.31 | 0.149 | 2 | 5 | 7 |
| 6.7 | Band 3 | 3.43 | 0.161 | 2 | 6 | 8 |

| SeptiScore range (midpoint) | RAPID Interpretation Band | Likelihood ratio (positive) | Likelihood ratio (negative) | No. of BC+ Sepsis patients | No. of SIRS patients | Total No. patients |
|---|---|---|---|---|---|---|
| 6.8 | Band 3 | 3.61 | 0.172 | 2 | 7 | 9 |
| 6.9 | Band 3 | 3.85 | 0.182 | 3 | 2 | 5 |
| 7 | Band 3 | 3.87 | 0.201 | 4 | 6 | 10 |
| 7.1 | Band 3 | 4.06 | 0.225 | 3 | 7 | 10 |
| 7.2 | Band 3 | 4.37 | 0.240 | 3 | 6 | 9 |
| 7.3 | Band 3 | 4.68 | 0.255 | 0 | 4 | 4 |
| 7.4 | Band 4 | 4.99 | 0.252 | 3 | 2 | 5 |
| 7.5 | Band 4 | 5.05 | 0.269 | 8 | 1 | 9 |
| 7.6 | Band 4 | 4.85 | 0.320 | 1 | 1 | 2 |
| 7.7 | Band 4 | 4.89 | 0.325 | 4 | 9 | 13 |
| 7.8 | Band 4 | 5.64 | 0.341 | 2 | 0 | 2 |
| 7.9 | Band 4 | 5.55 | 0.354 | 4 | 4 | 8 |
| 8 | Band 4 | 5.86 | 0.374 | 5 | 0 | 5 |
| 8.1 | Band 4 | 5.63 | 0.405 | 1 | 1 | 2 |
| 8.2 | Band 4 | 5.71 | 0.410 | 9 | 2 | 11 |
| 8.3 | Band 4 | 5.53 | 0.462 | 0 | 5 | 5 |
| 8.4 | Band 4 | 6.29 | 0.456 | 1 | 2 | 3 |
| 8.5 | Band 4 | 6.6 | 0.459 | 2 | 1 | 3 |
| 8.6 | Band 4 | 6.67 | 0.469 | 3 | 0 | 3 |
| 8.7 | Band 4 | 6.48 | 0.487 | 2 | 3 | 5 |
| 8.8 | Band 4 | 6.99 | 0.495 | 5 | 2 | 7 |
| 8.9 | Band 4 | 7.12 | 0.522 | 3 | 4 | 7 |
| 9 | Band 4 | 8.04 | 0.533 | 2 | 0 | 2 |
| 9.1 | Band 4 | 7.87 | 0.545 | 0 | 1 | 1 |
| 9.2 | Band 4 | 8.21 | 0.543 | 4 | 0 | 4 |
| 9.3 | Band 4 | 7.84 | 0.566 | 4 | 1 | 5 |
| 9.4 | Band 4 | 7.82 | 0.588 | 10 | 3 | 13 |
| 9.5 | Band 4 | 7.95 | 0.640 | 5 | 1 | 6 |
| 9.6-9.9 (mid 9.7) | Band 4 | 8.4 | 0.678 | 10 | 6 | 16 |
| 10.0-10.9 (mid 10.4) | Band 4 | 11.18 | 0.779 | 27 | 6 | 33 |
| 11.0-15.0 (mid 12.5) | Band 4 | 20.16 | 0.929 | 30 | 6 | 36 |
| | | | | **184** | **386** | **570** |

**Supplement S4:** Numbers of SIRS & sepsis patients per SeptiScore interval, in the LR analysis of BC(-) sepsis vs. SIRS. The dataset for this analysis consists of N=605 patients. Likelihood ratio estimates at the extreme ends of the SeptiScore distribution are based on wide SeptiScore intervals to allow sufficient numbers of SIRS & sepsis patients per interval for accurate LR estimation.

| SeptiScore range (midpoint) | RAPID Interpretation Band | Likelihood ratio (positive) | Likelihood ratio (negative) | No. of BC-Sepsis patients | No. of SIRS patients | Total No. patients |
|---|---|---|---|---|---|---|
| 1.8-3.3 (mid 2.7) | Band 1 | 1.02 | 0.031 | 1 | 15 | 16 |
| 3.4 | Band 1 | 1.04 | 0.118 | 0 | 5 | 5 |
| 3.5 | Band 1 | 1.05 | 0.088 | 0 | 6 | 6 |
| 3.6 | Band 1 | 1.07 | 0.068 | 0 | 3 | 3 |
| 3.7 | Band 1 | 1.08 | 0.061 | 1 | 4 | 5 |
| 3.8 | Band 1 | 1.08 | 0.107 | 0 | 11 | 11 |
| 3.9 | Band 1 | 1.12 | 0.080 | 1 | 1 | 2 |
| 4 | Band 1 | 1.12 | 0.118 | 1 | 4 | 5 |
| 4.1 | Band 1 | 1.12 | 0.144 | 2 | 14 | 16 |
| 4.2 | Band 1 | 1.16 | 0.168 | 0 | 10 | 10 |
| 4.3 | Band 1 | 1.2 | 0.145 | 0 | 7 | 7 |
| 4.4 | Band 1 | 1.23 | 0.132 | 1 | 8 | 9 |
| 4.5 | Band 1 | 1.25 | 0.140 | 2 | 4 | 6 |
| 4.6 | Band 1 | 1.26 | 0.172 | 2 | 12 | 14 |
| 4.7 | Band 1 | 1.3 | 0.186 | 0 | 13 | 13 |
| 4.8 | Band 1 | 1.36 | 0.166 | 0 | 10 | 10 |
| 4.9 | Band 1 | 1.42 | 0.153 | 2 | 12 | 14 |
| 5 | Band 2 | 1.47 | 0.165 | 0 | 8 | 8 |
| 5.1 | Band 2 | 1.52 | 0.156 | 1 | 10 | 11 |
| 5.2 | Band 2 | 1.58 | 0.157 | 1 | 16 | 17 |
| 5.3 | Band 2 | 1.69 | 0.153 | 1 | 8 | 9 |
| 5.4 | Band 2 | 1.75 | 0.156 | 1 | 9 | 10 |
| 5.5 | Band 2 | 1.82 | 0.158 | 2 | 9 | 11 |
| 5.6 | Band 2 | 1.89 | 0.168 | 1 | 9 | 10 |
| 5.7 | Band 2 | 1.97 | 0.169 | 1 | 10 | 11 |
| 5.8 | Band 2 | 2.08 | 0.170 | 1 | 9 | 10 |
| 5.9 | Band 2 | 2.18 | 0.171 | 3 | 10 | 13 |

| SeptiScore range (midpoint) | RAPID Interpretation Band | Likelihood ratio (positive) | Likelihood ratio (negative) | No. of BC-Sepsis patients | No. of SIRS patients | Total No. patients |
|---|---|---|---|---|---|---|
| 6 | Band 2 | 2.29 | 0.186 | 4 | 6 | 10 |
| 6.1 | Band 2 | 2.34 | 0.210 | 2 | 13 | 15 |
| 6.2 | Band 3 | 2.55 | 0.213 | 0 | 6 | 6 |
| 6.3 | Band 3 | 2.67 | 0.209 | 3 | 7 | 10 |
| 6.4 | Band 3 | 2.79 | 0.223 | 1 | 5 | 6 |
| 6.5 | Band 3 | 2.9 | 0.225 | 5 | 8 | 13 |
| 6.6 | Band 3 | 3.03 | 0.250 | 2 | 5 | 7 |
| 6.7 | Band 3 | 3.15 | 0.258 | 1 | 6 | 7 |
| 6.8 | Band 3 | 3.34 | 0.259 | 2 | 7 | 9 |
| 6.9 | Band 3 | 3.57 | 0.264 | 7 | 2 | 9 |
| 7 | Band 3 | 3.5 | 0.303 | 4 | 6 | 10 |
| 7.1 | Band 3 | 3.68 | 0.320 | 4 | 7 | 11 |
| 7.2 | Band 3 | 3.95 | 0.336 | 0 | 6 | 6 |
| 7.3 | Band 3 | 4.31 | 0.329 | 4 | 4 | 8 |
| 7.4 | Band 4 | 4.48 | 0.347 | 4 | 2 | 6 |
| 7.5 | Band 4 | 4.51 | 0.367 | 5 | 1 | 6 |
| 7.6 | Band 4 | 4.44 | 0.392 | 4 | 1 | 5 |
| 7.7 | Band 4 | 4.39 | 0.413 | 5 | 9 | 14 |
| 7.8 | Band 4 | 5.03 | 0.428 | 5 | 0 | 5 |
| 7.9 | Band 4 | 4.85 | 0.454 | 2 | 4 | 6 |
| 8 | Band 4 | 5.21 | 0.459 | 4 | 0 | 4 |
| 8.1 | Band 4 | 5.05 | 0.479 | 4 | 1 | 5 |
| 8.2 | Band 4 | 5 | 0.498 | 2 | 2 | 4 |
| 8.3 | Band 4 | 5.16 | 0.506 | 2 | 5 | 7 |
| 8.4 | Band 4 | 5.78 | 0.509 | 4 | 2 | 6 |
| 8.5 | Band 4 | 5.91 | 0.526 | 2 | 1 | 3 |
| 8.6 | Band 4 | 5.98 | 0.534 | 4 | 0 | 4 |
| 8.7 | Band 4 | 5.77 | 0.554 | 6 | 3 | 9 |
| 8.8 | Band 4 | 5.99 | 0.579 | 4 | 2 | 6 |
| 8.9 | Band 4 | 6.17 | 0.596 | 4 | 4 | 8 |
| 9 | Band 4 | 6.9 | 0.609 | 2 | 0 | 2 |
| 9.1 | Band 4 | 6.76 | 0.618 | 4 | 1 | 5 |
| 9.2 | Band 4 | 6.74 | 0.636 | 3 | 0 | 3 |

| SeptiScore range (midpoint) | RAPID Interpretation Band | Likelihood ratio (positive) | Likelihood ratio (negative) | No. of BC-Sepsis patients | No. of SIRS patients | Total No. patients |
|---|---|---|---|---|---|---|
| 9.3 | Band 4 | 6.51 | 0.651 | 5 | 1 | 6 |
| 9.4 | Band 4 | 6.41 | 0.673 | 7 | 3 | 10 |
| 9.5 | Band 4 | 6.77 | 0.701 | 2 | 1 | 3 |
| 9.6-9.9 (mid 9.7) | Band 4 | 7.66 | 0.718 | 10 | 6 | 16 |
| 10.0-10.9 (mid 10.4) | Band 4 | 10.85 | 0.784 | 24 | 6 | 30 |
| 11.0-15.0 (mid 12.5) | Band 4 | 14.67 | 0.935 | 37 | 6 | 43 |
| | | | | **219** | **386** | **605** |

**Supplement S5: Further analysis of J-shaped Curve in Figure 5C**

In Figure 5C, the $LR^-$ analysis for the BC(+) versus SIRS comparison, we observe a distinctive J-shaped curve with a minimum near 5.0 (Figure 5C, blue points). To understand this behavior, we examined the components of the $LR^-$ formula, LR- = (FN/TN) x (TN+FP) / (TP+FN). The second term is constant, because TN+FP equals the total number of negative calls. The third term is also constant, because TP+FN equals the total number of positive calls. Consequently, all variation in $LR^-$ must arise from the FN/TN term. When FN/TN is plotted as a function of SeptiScore, its curve closely mirrors the $LR^-$ curve (Figure 5C, orange points, reproduced below).

The "J" shape of the curve between the limits of 5.0 and 3.3 easily explained, as follows. The second-lowest BC(+) sepsis patient has SeptiScore = 5.0, and the lowest BC(+) sepsis patient has SeptiScore 3.3. As the cutoff is decreased below 5.0 toward 3.3, the numerator of FN/TN remains constant at FN=1 (because there is only the single BC(+) sepsis patient remaining, while the denominator TN (equal to the number of true negatives below the cutoff) continues to decrease. Thus, the ratio FN/TN increases monotonically as the cutoff is lowered.

**Figure 5C in the main text:**

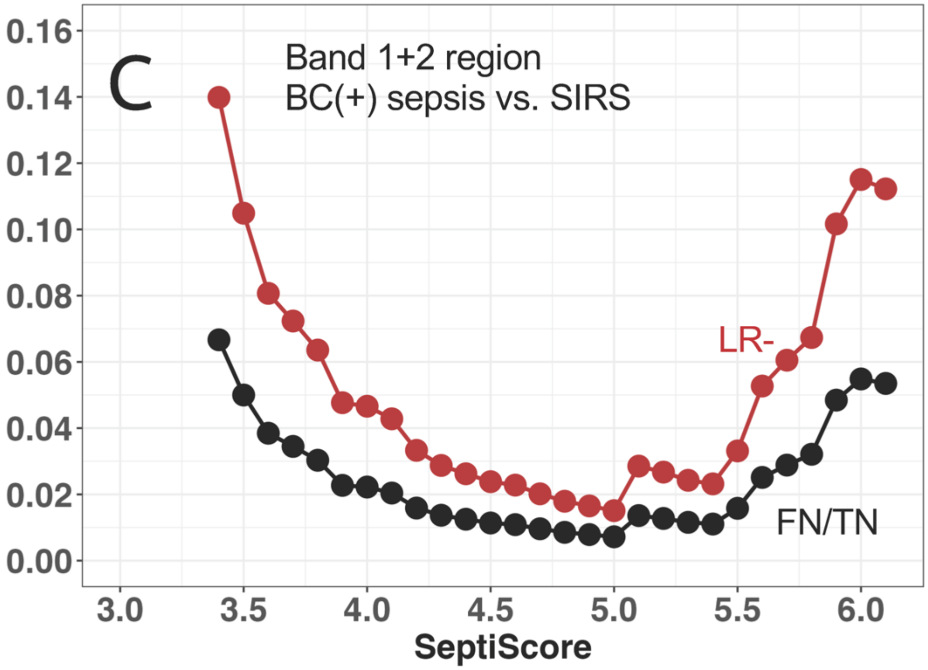
C
Band 1+2 region
BC(+) sepsis vs. SIRS
LR-
FN/TN
0.16
0.14
0.12
0.10
0.08
0.06
0.04
0.02
0.00
3.0
3.5
4.0
4.5
5.0
5.5
6.0
SeptiScore

**Supplement S6: Comparative Estimates of Sepsis pre-test Probability**

In the acute care setting considered here, the relevant time horizon is very short—approximately ±1 day around the attending physician's decision to initiate or withhold a sepsis bundle (including intravenous antibiotics and microbiological cultures) in response to a suspected new sepsis event. Over such a narrow interval, the pre-test probability can be reasonably approximated by the short-interval incidence of sepsis, as disease status evolves rapidly and the distinction between point prevalence and incidence becomes negligible at this time scale. Accordingly, for the purposes of the present analysis, we assume the pre-test probability to be the cohort-level short-interval incidence of sepsis, as determined by expert adjudication.

We wish to place the estimated pre-test probability of sepsis (50% ± 2.5%) into the broader context of what might be expected on the basis of other studies from the literature. In the Intensive Care over Nations (ICON) audit (Sakr et al., 2018), estimates of sepsis incidence in the ICU vary widely, with an average incidence of 29.5% and a range of 13.6% to 39.3% depending upon region. This variation most likely depends on multiple factors, as enumerated in the following table.

**Table S5:** Factors reported to affect the estimates of sepsis incidence (pre-test probability)

| **Influencing Factor** | **Likely effect** | **Reference** |
|---|---|---|
| Framework definition: sepsis-2 vs. sepsis-3 | Sepsis-3 more restrictive, leading to fewer patients called septic, therefore lower incidence rate | Shankar-Hari et al. (2017) [28]<br>Hammond et al. (2022) [29] |
| ICD code-based vs. chart-based review | Chart review more accurate, but more resource intensive. | Mariansdatter et al. (2016) [30] |
| Cohort specific factor: comorbidities | Sepsis incidence increases with particular comorbidities: COPD, heart failure, renal failure, cirrhosis, immunosuppression, HIV-1 infection | Sakr et al. (2018) [23] |
| Cohort specific factor: geographic location | Higher infectious disease burden in some locations leads to higher sepsis incidence | Sakr et al. (2018) [23]<br>GBD (2021) [31] |
| Cohort specific factor: race/ethnicity | Higher incidence of severe sepsis reported for Black, as opposed to Hispanic or White (root cause not identified) | Barnato et al. (2008) [32] |
